# Clinical activity of MAPK targeted therapies in patients with non-V600 BRAF mutant tumors

**DOI:** 10.1101/2022.02.17.22271120

**Authors:** Matthew Dankner, Yifan Wang, Rouhi Fazelzad, Benny Johnson, Caroline A. Nebhan, Ibiayi Dagogo-Jack, Nathaniel J. Myall, Georg Richtig, Jillian W. P Bracht, Marco Gerlinger, Eiji Shinozaki, Takayuki Yoshino, Daisuke Kotani, Jason R. Fangusaro, Oliver Gautschi, Julien Mazieres, Jeffrey A. Sosman, Scott Kopetz, Vivek Subbiah, Michael A. Davies, Anna Groover, Ryan J. Sullivan, Keith T. Flaherty, Douglas B. Johnson, Andrea Benedetti, David W. Cescon, Anna Spreafico, George Zogopoulos, April A. N. Rose

## Abstract

**Purpose:** Non-V600 mutations comprise approximately 35% of all BRAF mutations in cancer. Many of these mutations have been identified as oncogenic drivers in a wide array of cancer types and can be classified into three Classes according to molecular characteristics. Consensus treatment strategies for Class 2 and 3 BRAF mutations have not yet been established.

**Methods:** We performed a systematic review and meta-analysis of individual patient data to assess treatment outcomes with FDA-approved mitogen activated protein kinase pathway (MAPK) targeted therapy according to BRAF Class, cancer type and MAPK targeted therapy type. A search was conducted on literature from 2010-2021. Individual patient data was collected and analyzed from published reports of patients with cancer harboring Class 2 or 3 BRAF mutations and who received MAPK targeted therapy with available treatment response data. Co-primary outcomes were response rate (RR) and progression-free survival (PFS).

**Results:** 18167 studies were screened, identifying 80 studies with 238 patients that met inclusion criteria. This included 167 patients with Class 2 and 71 patients with Class 3 BRAF mutations. Overall, 77 patients achieved a treatment response. In both univariate and multivariable analyses, RR and PFS were higher among patients with Class 2 compared to Class 3 mutations, findings that remain when analyses are restricted to patients with melanoma or lung primary cancers. MEK +/- BRAF inhibitors demonstrated greater clinical activity in Class 2 compared to Class 3 BRAF mutant tumors than BRAF or EGFR inhibitors.

**Conclusions:** This meta-analysis suggests that MAPK targeted therapies have clinical activity in some Class 2 and 3 BRAF mutant cancers. BRAF Class may dictate responsiveness to current and emerging treatment strategies, particularly in metastatic melanoma and lung cancers. Together, this analysis provides clinical validation of predictions made based on a mutation classification system established in the preclinical literature. Further evaluation with prospective clinical trials is needed for this population.

## Introduction

BRAF is among the most commonly mutated genes in human cancer [1]. BRAF is most frequently mutated at codon V600, resulting in enhanced activation of the downstream mitogen activated protein kinase (MAPK) pathway [1]. Randomized clinical trials investigating targeted therapy (MAPK TT) strategies using BRAF, MEK, BRAF + MEK, and BRAF ± MEK + EGFR inhibitors have yielded response rates of >50% in patients with BRAF V600 mutant tumors [2–9]. Several of these trials have demonstrated overall survival benefit for these therapeutic strategies [2, 4, 9–11]. As a result, MAPK TT are now standard of care treatments for patients with BRAF V600 mutant melanoma, lung cancer, and colorectal cancer [12–14].

Approximately 35% of all BRAF mutations occur outside the V600 codon [1, 15]. In addition to missense mutations, recurring oncogenic BRAF fusions and in-frame deletions have also been described [16–18]. Seminal preclinical work by Wan *et al.* demonstrated that many non-V600 mutations are oncogenic and result in altered kinase activity [19]. More recently, differences in dimerization requirement and RAS dependency in frequently identified non-V600 BRAF mutations have been described by Yao *et al.* [20, 21]. The combination of these molecular data has led to a classification scheme for BRAF alterations [15, 21]. Wild-type BRAF signals as RAS-dependent dimers, and Class 1 BRAF mutants are comprised of V600-mutations, which signal as constitutively active monomers in a RAS-independent manner [22, 23]. Class 2 BRAF mutations form kinase-activating RAS-independent dimers [20], and Class 3 BRAF mutations have impaired kinase activity but signal as RAS-dependent dimers, primarily by forming heterodimers with CRAF [21].

The sensitivity of Class 2 and Class 3 BRAF mutant tumors to MAPK TT is unclear. There are preclinical data that support the use of MEK inhibitors +/- BRAF inhibitors in tumors with Class 2 or 3 mutations [24–27]. Due to the dependency on RAS activation, receptor tyrosine kinase (RTK) inhibitors +/- MEK inhibitors have been proposed as a viable therapeutic strategy for Class 3 BRAF mutant tumors [21]. However, preclinical evidence also suggests that non-V600 BRAF mutations may be less sensitive to BRAF + MEK inhibition than Class 1 mutant tumors [20, 21]. Recently, two single-arm Phase II trials have reported response rates for the MEK inhibitor, trametinib, in melanoma patients (33%, n=9) and in a tumor-agnostic cohort of patients (3%, n=32) with non-V600 BRAF mutations [28, 29]. However, a multitude of case reports and case series in different cancer types have demonstrated that subsets of non-V600 BRAF mutant tumors may indeed be sensitive to these FDA-approved agents [1, 24].

There are currently no data from randomized controlled trials to guide targeted therapy treatment decisions in cancers with Class 2/3 BRAF mutations. As such, there is clinical equipoise regarding the best targeted treatment strategy for patients whose tumors express these important driver oncogenes. When standard treatment options have been exhausted, many oncologists will provide off-label MAPK targeted therapies to these patients. Therefore, to establish a reference cohort that could help guide treatment decisions and inform future clinical trial design, we sought to compile and synthesize all available clinical evidence in the medical literature wherein Class 2 or 3 BRAF mutant tumors were treated with MAPK TT.

## Methods

### Search Strategy

A literature search was conducted of studies published from January 2010 to September 2021 in the following databases: Medline ALL (Medline and Medline Epub Ahead of print and In-Process & Other Non-Indexed Citations), Embase, Cochrane Central Register of Controlled Trials, Cochrane Database of Systematic Reviews, all from the OvidSP platform, and Web of Science from Clarivate Analytics. Where available, both controlled vocabulary terms and text words were used (Appendix 1). There were no language or study design restrictions. Published conference abstracts were included. The reference lists of included studies were scanned to identify additional relevant studies. The American Association for Cancer Research (AACR), the American Society of Clinical Oncology (ASCO) and the European Society of Medical Oncology (ESMO) conference proceedings were searched to identify any relevant conference abstracts. Additional publications and/or data identified by the authors outside of the search were added to the systematic review when applicable. The study protocol was prospectively uploaded to PROSPERO (ID: CRD42020218141) and followed the preferred reporting items for systematic reviews and meta-analyses (PRISMA) guidelines [30].

Abstracts were screened by two independent reviewers using Covidence software (www.covidence.org). Conflicts were resolved with internal discussion between the two reviewers and in the case of a lasting conflict, by a third reviewer. Response data and patient demographics were extracted by two independent reviewers. After data was extracted from all included publications, missing data was identified and requested from the original authors with up to two separate email prompts >7 days apart.

### Inclusion and exclusion criteria

Inclusion criteria were: published reports of adult patients with cancer with individual patient data describing 1) a Class 2 or Class 3 BRAF mutation, 2) treatment with FDA-approved MAPK TT including inhibitors of EGFR, BRAF monomers, or MEK, and 3) availability of treatment response data. Exclusion criteria were: the presence of a concomitant BRAF V600E/K mutation, pediatric patients, concurrent systemic non-MAPK TT (such as chemotherapy, immunotherapy, PI3K or CDK4/6 targeted agents).

### Primary and secondary outcomes

The co-primary outcomes were overall treatment response rate (RR) and progression-free survival (PFS). When appropriate response criteria were used (RECIST), patients with partial response (PR) or complete response (CR) were considered to have had a treatment response and those with stable disease (SD) or (PD) were considered non-responders [31]. When RECIST criteria were not used, response was recorded based on the primary paper’s author’s assessment of response or calculated from tumor measurements on CT or MRI provided in the text. For PFS analysis, patients were censored if there was no indication of progression or death at the time of last follow-up.

### Quality (risk of bias) assessment

To assess the methodological quality of individual studies included in the study, we used a previously described tool that is adapted for evaluation of case reports and case series. The tool includes 5 items that are derived from the Newcastle-Ottawa scale [32]. These 5 items examine the selection and representativeness of cases and the ascertainment of outcome and exposure, with each item scored one point if the information was specifically reported. We deemed the study to be of good quality (low risk of bias) when all 5 criteria were met, of moderate quality when 4 criteria were met, and of poor quality (high risk of bias) when ≤3 criteria were fulfilled.

### Statistical Analyses

We performed one-stage meta-analyses of pooled individual patient level data from all included studies. Hazard ratio was used as the parameter of interest for PFS, and odds ratios (OR) was used as the parameter of interest for response. A multi-level mixed effects logistic regression model, incorporating individual study as a random effect, was used to estimate the odds ratios of responses between groups and its associated 95% confidence interval. Multivariable logistic regression models, incorporating study type, cancer type and therapy type were used to estimate adjusted odds ratios. For multivariable analysis of treatment response, all study key variables that were available for all patients were incorporated into the initial multivariable model. These included: cancer type, BRAF mutation class, therapy type, geographic location, study type, and response criteria. The final multivariable models for PFS included only those variables that were associated with P<0.05. To analyze progression-free survival, a shared frailty Cox-regression model was used to account for heterogeneity across studies for all primary analyses. For multivariable analysis of progression-free survival, all study key variables were incorporated into the initial multivariable model. These included: age, sex, cancer type, BRAF mutation class, therapy type, geographic location, study type, and response criteria. We performed backward selection to identify potentially significant variables. The final multivariable models for PFS included only those variables that were associated with P<0.05. Survival curves were visualized with the Kaplan-Meier method and the log-rank was used to test differences in survival between populations. Statistical analyses were performed with STATA v13.

## Results

### Characteristics of included studies and patients

We identified 18,167 potentially eligible articles in our search. After removing ineligible articles and adding additional studies from the author’s files, a total of 80 articles were included in the review (Appendix 2), comprising a total of 238 cancer patients with Class 2 or Class 3 non-V600 BRAF mutations who were treated with MAPK targeted therapy (Figure 1). The number of studies reporting results of MAPK inhibitor treatment outcomes in patients with tumors harboring non-V600 BRAF mutations has increased substantially over the past decade (Supplemental Figure S1). A detailed description of the different MAPK targeted therapy treatment regimens used for patients in the study is presented in Supplemental Table S1. We also performed a risk of bias assessment for all studies included in the meta-analysis on a 5-point scale (Supplemental Figure S2).

**Figure 1:**
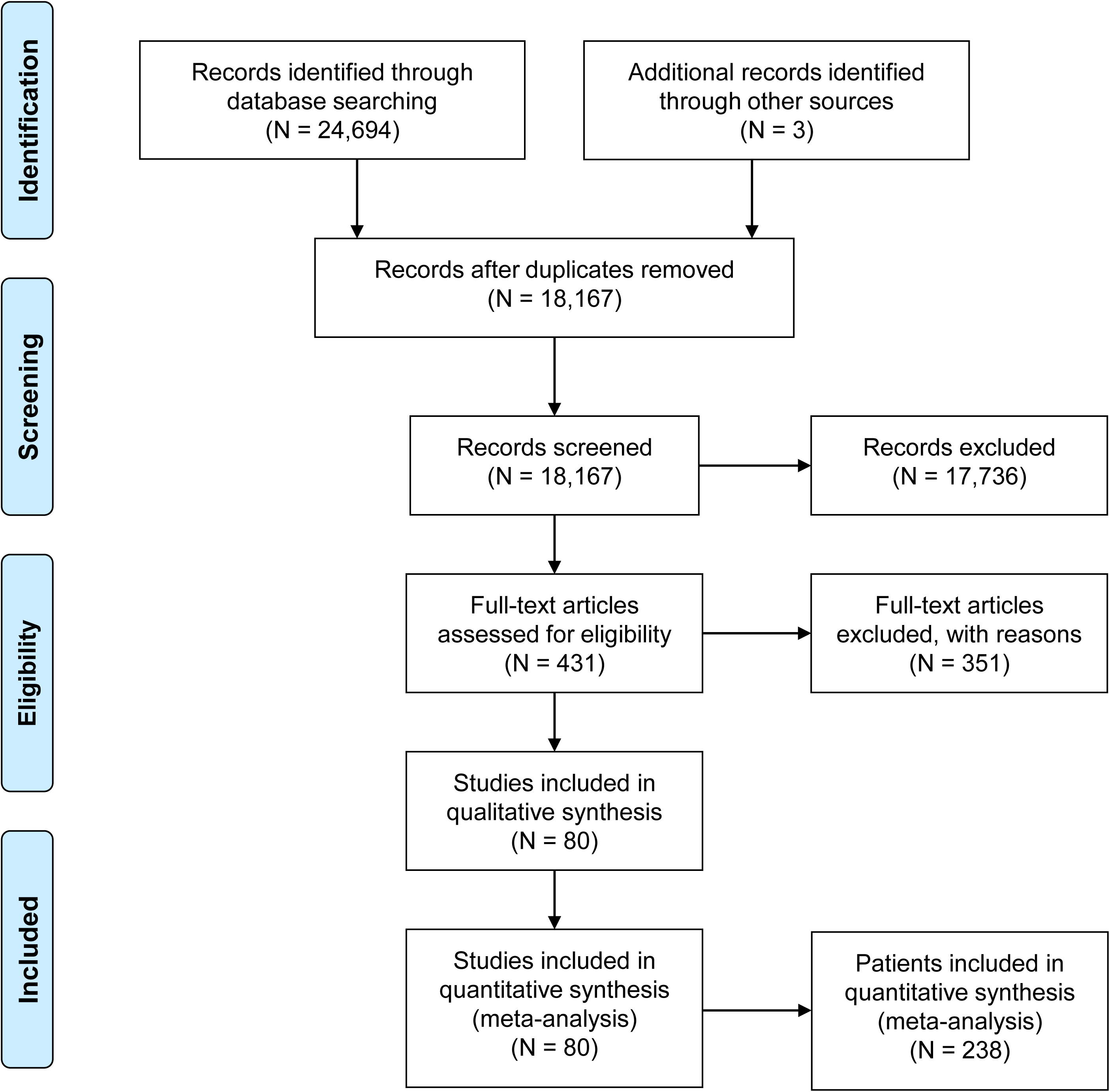
PRISMA diagram demonstrating search and inclusion of studies for meta-analysis.

**Table 1:** Individual patient characteristics

|  | Entire Cohort | Class 2 | Class 3 | P (Fischer's Exact) | Pearson's Chi-Squared |
| --- | --- | --- | --- | --- | --- |
| *Entire Cohort* | 238 | 167 (70%) | 71 (30%) |  |  |
| *Study Characteristics* |  |  |  |  |  |
| **Study Type** |  |  |  |  |  |
| Prospective | 77 (32%) | 37 (48%) | 40 (52%) | **<0.001** |  |
| Retrospective | 161 (68%) | 130 (81%) | 31 (19%) |  |  |
| **Response Criteria** |  |  |  |  |  |
| RECIST | 112 (47%) | 68 (61%) | 44 (39%) | **0.003** |  |
| non-RECIST | 126 (53%) | 99 (79%) | 27 (21%) |  |  |
| **Geographic Location** |  |  |  |  |  |
| North America | 123 (52%) | 75 (61%) | 48 (39%) | **0.002** | **12.91, P=0.005** |
| Europe | 93 (39%) | 75 (81%) | 18 (19%) | **0.006** |  |
| Asia | 15 (6%) | 10 (67%) | 5 (33%) | 0.774 |  |
| Australia | 7 (3%) | 7 (100%) | 0 (0%) | 0.107 |  |
| *Patient & Treatment Characteristics* |  |  |  |  |  |
| **Gender** |  |  |  |  |  |
| Male | 70 (29%) | 50 (71%) | 20 (29%) | 0.323 |  |
| Female | 48 (20%) | 30 (63%) | 18 (37%) |  |  |
| Unknown | 120 (50%) | 87 (73%) | 33 (27%) |  |  |
| **Age** |  |  |  |  |  |
| >= 65 | 49 (21%) | 31 (63%) | 18 (32%) | 0.311 |  |
| < 65 | 62 (26%) | 45 (73%) | 17 (27%) |  |  |
| Unknown | 127 (53%) | 91 (72%) | 36 (28%) |  |  |
| **Cancer Type** |  |  |  |  |  |
| Colorectal Cancer | 12 (5%) | 5 (42%) | 7 (58%) | **0.046** | **24.70, P<0.001** |
| Lung Cancer | 58 (24%) | 32 (55%) | 26 (45%) | **0.005** |  |
| Melanoma | 117 (49%) | 99 (85%) | 18 (15%) | **<0.001** |  |
| Other | 51 (21%) | 31 (61%) | 20 (39%) | 0.084 |  |
| **Therapy Type** |  |  |  |  |  |
| BRAFi | 79 (33%) | 57 (72%) | 22 (28%) | 0.88 | **8.6738, P=0.034** |
| MEKi | 85 (36%) | 52 (61%) | 33 (39%) | **0.041** |  |
| BRAFi + MEKi | 63 (26%) | 52 (83%) | 11 (17%) | **0.015** |  |
| EGFRi | 11 (5%) | 6 (55%) | 5 (45%) | 0.311 |  |
| **RAS Mutation** |  |  |  |  |  |
| Present | 14 (6%) | 3 (21%) | 11 (79%) | **0.004** |  |
| Absent | 122 (51%) | 77 (63%) | 45 (37%) |  |  |
| Unknown | 102 (43%) | 87 (85%) | 15 (15%) |  |  |
| **Table 1: Individual patient characteristics** |  |  |  |  |  |

Among the 238 patients included in this study, there were 167 patients with Class 2 and 71 patients with Class 3 BRAF mutations (Table 1). The details of mutations categorized as Class 2 and 3 are described in Supplemental Table S2.

**Table 2:** Overall response rates associated with clinical variables. Odds ratios, 95% confidence intervals and P-values calculated with a multilevel mixed-effects logistic regression model with article as the random-effects variable.

|  | # of Patients | # of Patients with Response | Response Rate | Univariate Odds Ratio | Univariate 95% CI | Univariate P Value | Adjusted Odds Ratio | Multivariate 95% CI | Adjusted P Value |
| --- | --- | --- | --- | --- | --- | --- | --- | --- | --- |
| *Entire Cohort* | 238 | 77 | 32.4% |  |  |  |  |  |  |
| *Study Characteristics* |  |  |  |  |  |  |  |  |  |
| **Study Type** |  |  |  |  |  |  |  |  |  |
| Prospective | 77 | 10 | 13.0% | 0.089 | .0165-.478 | **0.005** |  |  |  |
| Retrospective | 161 | 67 | 41.6% |  |  |  |  |  |  |
| **Response Criteria** |  |  |  |  |  |  |  |  |  |
| RECIST | 112 | 24 | 21.4% | 0.121 | .0285-.516 | **0.004** | 0.142 | 0.0402-0.499 | **0.002** |
| non-RECIST | 126 | 53 | 42.1% |  |  |  |  |  |  |
| **Geographic Location** |  |  |  |  |  |  |  |  |  |
| North America | 123 | 38 | 30.9% | 0.929 | 0.232-3.711 | 0.917 |  |  |  |
| Europe | 93 | 29 | 31.2% | 1.417 | 0.342-5.880 | 0.631 |  |  |  |
| Asia | 15 | 6 | 40.0% | 0.510 | 0.0494-5.255 | 0.571 |  |  |  |
| Australia | 7 | 4 | 57.1% | 0.982 | 0.0265-36.413 | 0.992 |  |  |  |
| *Patient & Treatment Characteristics* |  |  |  |  |  |  |  |  |  |
| **Gender** |  |  |  |  |  |  |  |  |  |
| Male | 70 | 32 | 45.7% | 0.805 | 0.255-2.541 | 0.711 |  |  |  |
| Female | 48 | 25 | 52.1% |  |  |  |  |  |  |
| Unknown | 120 | 20 | 16.7% |  |  |  |  |  |  |
| **Age** |  |  |  |  |  |  |  |  |  |
| >= 65 | 49 | 26 | 53.1% | 1.139 | 0.362-3.583 | 0.824 |  |  |  |
| < 65 | 62 | 30 | 48.4% |  |  |  |  |  |  |
| Unknown | 127 | 21 | 16.5% |  |  |  |  |  |  |
| **Cancer Type** |  |  |  |  |  |  |  |  |  |
| Colorectal Cancer | 12 | 2 | 16.7% | 0.162 | 0.161-1.621 | 0.121 |  |  |  |
| Lung Cancer | 58 | 16 | 27.6% | 0.720 | 0.190-2.726 | 0.628 |  |  |  |
| Melanoma | 117 | 41 | 35.0% | 2.147 | 0.569-8.106 | 0.26 |  |  |  |
| Other | 51 | 18 | 35.3% | 1.094 | 0.296-4.049 | 0.893 |  |  |  |
| **Therapy Type** |  |  |  |  |  |  |  |  |  |
| BRAF<sup>i</sup> | 77 | 9 | 11.7% | 0.080 | 0.0201-0.316 | **<0.001** |  |  |  |
| MEK<sup>i</sup> | 87 | 64 | 37.9% | 2.291 | 0.768-6.383 | 0.137 | 9.735 | 2.567-36.914 | **0.001** |
| BRAF<sup>i</sup> + MEK<sup>i</sup> | 63 | 32 | 50.8% | 4.182 | 1.544-11.324 | **0.005** | 10.947 | 3.118-38.432 | **<0.001** |
| EGFR<sup>i</sup> | 11 | 3 | 27.3% | 0.191 | 0.0132-2.780 | 0.226 |  |  |  |
| **RAS Mutation** |  |  |  |  |  |  |  |  |  |
| Present | 14 | 1 | 7.1% | 0.056 | 0.0028-1.11 | 0.058 |  |  |  |
| Absent | 122 | 47 | 38.5% |  |  |  |  |  |  |
| Unknown | 102 | 29 | 28.4% |  |  |  |  |  |  |
| **BRAF Mutation Class** |  |  |  |  |  |  |  |  |  |
| 2 | 167 | 68 | 40.7% | 5.120 | 1.847-14.167 | **0.002** | 5.836 | 2.015-16.902 | **0.001** |
| 3 | 71 | 9 | 12.7% |  |  |  |  |  |  |

### Characteristics associated with MAPK inhibitor response and progression-free survival by BRAF Class and primary tumor type

In the entire population, 77 out of 238 patients (32%) experienced a treatment response (Table 2). The treatment response rate (RR) differed according to whether tumors had a Class 2 or Class 3 BRAF mutation (41% vs. 13%, univariable OR 5.12, P=0.002; Table 2). We next compared the impact of BRAF mutation Class on treatment response within each primary tumor type. Class 2 BRAF mutant tumors demonstrated higher response rates than Class 3 mutants independently in lung, melanoma, and ‘other’ primaries (P=0.018, P=0.029 and P=0.018, respectively; Figure 2A). Among those with Class 2 BRAF mutations, MAPK targeted therapy response rates were highest in patients with “other” tumor types (48%) and lowest in colorectal cancer patients (20%) (Figure 2A). The group of 51 patients with ‘other’ tumor types had non-colorectal gastrointestinal (n=21), genitourinary (n=10), gynecological (n=5), hematopoietic (n=4), head and neck (n=4), breast (n=2), spindle cell neoplasms (n=1), low grade glioma (n=1) and unknown primary tumors (n=3). Among patients whose tumors harbored Class 3 mutations, response rates did not differ significantly according to primary tumor type (RR 11-15%) (Figure 2A).

**Figure 2:**
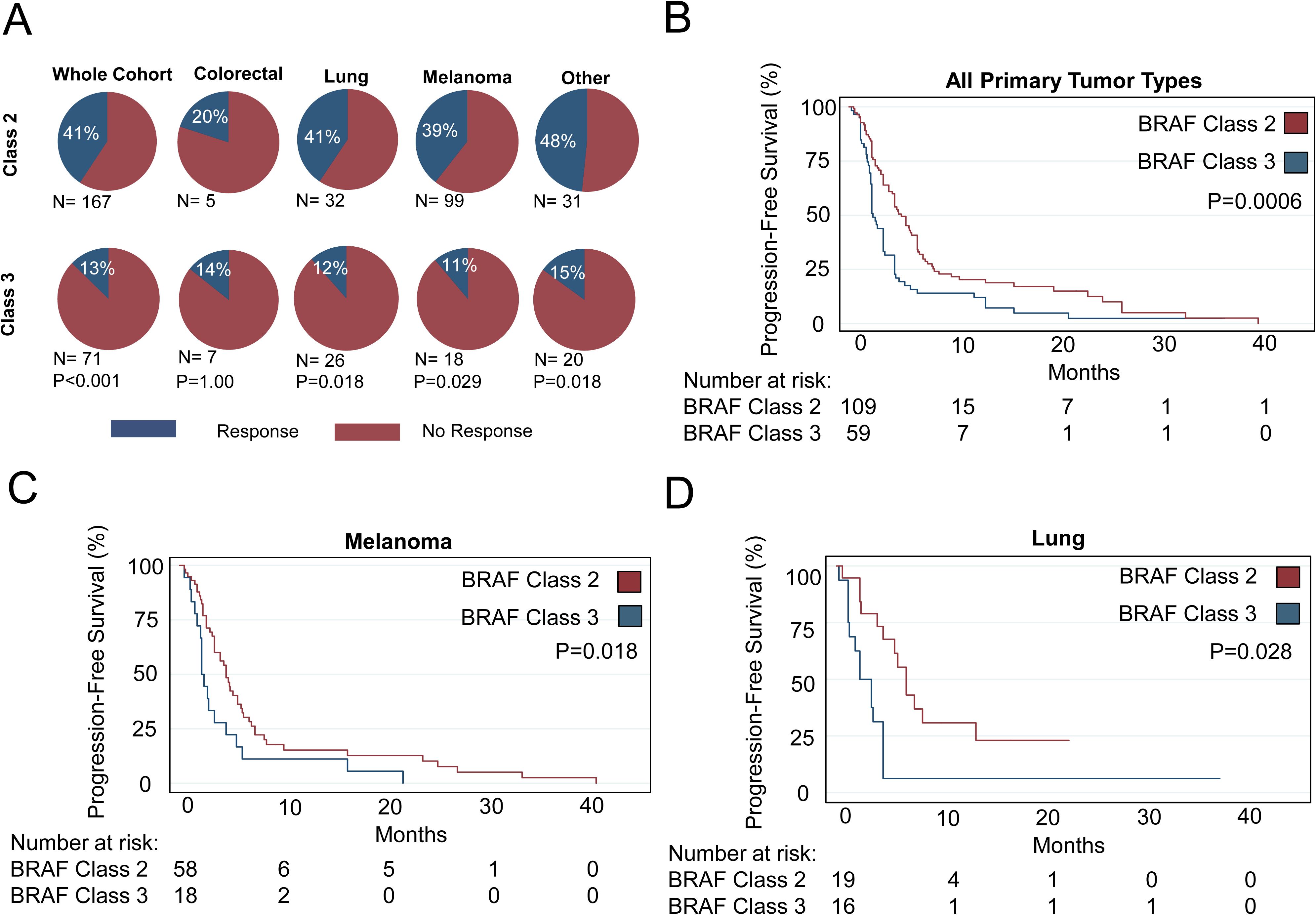
Relationship between BRAF Class and tumor type in the context of MAPK targeted therapy. A) Response rates to MAPK targeted therapy according to BRAF Class and primary cancer type. P-values calculated with Fischer’s Exact Test. B) Progression-free survival according to BRAF Class in the entire cohort and for melanoma (C) and lung (D) primary tumors. P-values calculated with Log-Rank test.

Data on progression-free survival (PFS) was available for 168 (71%) patients included in the study. Patients with Class 2 BRAF mutations (median PFS (mPFS) 4.6 months, hazard ratio (HR) 0.537, P=0.001) experienced longer PFS compared to patients with Class 3 mutations (mPFS 2.1 months) (Table 3, Figure 2B). The relationship between BRAF Class and PFS remained significant when we examined specific cancer subsets, including: metastatic melanoma (P=0.018) or lung cancer (P=0.028; Figure 2C and D, Supplemental Figure S3). When restricting our analyses to patients with RECIST-defined responses, from prospective datasets, or who were treated with only BRAF and/or MEK inhibitors, the differential PFS between Class 2 and Class 3 BRAF mutants remain (P=0.024, P=0.011 and P=0.002, respectively; Supplemental Figures S4 and S5).

**Table 3:** Progression-free survival associated with clinical variables. Hazard ratios, 95% confidence intervals and P-values calculated with a Cox proportional hazards model with article as the shared frailty variable.

|  | # of Patients with PFS data | Median PFS | Univariate Hazard Ratio | Univariate 95% CI | Univariate P Value | Multivariate Hazard Ratio | Multivariate 95% CI | Multivariate P Value |
| --- | --- | --- | --- | --- | --- | --- | --- | --- |
| *Study Characteristics* |  |  |  |  |  |  |  |  |
| **Study Type** |  |  |  |  |  |  |  |  |
| Prospective | 59 | 2.3 | 1.26 | 0.763-2.081 | 0.367 |  |  |  |
| Retrospective | 109 | 4 |  |  |  |  |  |  |
| **Response Criteria** |  |  |  |  |  |  |  |  |
| RECIST | 93 | 3 | 1.216 | 0.7883-1.872 | 0.378 |  |  |  |
| non-RECIST | 75 | 4 |  |  |  |  |  |  |
| **Geographic Location** |  |  |  |  |  |  |  |  |
| North America | 119 | 3 | 1.218 | 0.779-1.903 | 0.387 |  |  |  |
| Europe | 30 | 4 | 0.924 | 0.551-1.550 | 0.765 |  |  |  |
| Asia | 12 | 6 | 0.553 | 0.255-1.202 | 0.135 |  |  |  |
| Australia | 7 | 3.8 | 1.909 | 0.589-6.188 | 0.281 |  |  |  |
| *Patient & Treatment Characteristics* |  |  |  |  |  |  |  |  |
| **Gender** |  |  |  |  |  |  |  |  |
| Male | 66 | 3 | 1.122 | 0.711-1.771 | 0.62 |  |  |  |
| Female | 47 | 4.3 |  |  |  |  |  |  |
| **Age** |  |  |  |  |  |  |  |  |
| > / = 65 | 60 | 4 | 1.008 | 0.635-1.602 | 0.972 |  |  |  |
| < 65 | 49 | 4 |  |  |  |  |  |  |
| **Cancer Type** |  |  |  |  |  |  |  |  |
| Colorectal Cancer | 11 | 2.3 | 1.548 | 0.737-3.210 | 0.252 |  |  |  |
| Lung Cancer | 35 | 4 | 0.877 | 0.547-1.405 | 0.584 |  |  |  |
| Melanoma | 76 | 3.5 | 1.309 | 0.851-2.012 | 0.221 |  |  |  |
| Other | 46 | 4.1 | 0.776 | 0.492-1.225 | 0.276 |  |  |  |
| **Therapy Type** |  |  |  |  |  |  |  |  |
| BRAF<sup>i</sup> | 37 | 3.5 | 1.697 | 1.038-2.771 | **0.035** |  |  |  |
| MEK<sup>i</sup> or ERK<sup>i</sup> | 79 | 3 | 0.539 | 0.345-0.844 | **0.007** | 0.588 | 0.359-0.965 | **0.036** |
| BRAF<sup>i</sup> + MEK<sup>i</sup> | 42 | 5 | 0.646 | 0.408-1.022 | 0.062 | 0.462 | 0.266-0.801 | **0.006** |
| EGFR<sup>i</sup> | 10 | 2.8 | 0.669 | 0.801-3.685 | 0.164 |  |  |  |
| **RAS Mutation** |  |  |  |  |  |  |  |  |
| Present | 14 | 2 | 1.240 | 0.678-2.262 | 0.484 |  |  |  |
| Absent | 114 | 3 |  |  |  |  |  |  |
| **BRAF Mutation Class** |  |  |  |  |  |  |  |  |
| 2 | 109 | 4.6 | 0.537 | 0.366-0.788 | **0.001** | 0.544 | 0.375-0.789 | **0.001** |
| 3 | 59 | 2.1 |  |  |  |  |  |  |

### Characteristics associated with MAPK inhibitor response and progression-free survival by BRAF Class and treatment type

In Class 2 BRAF mutant tumors, the highest response rate was observed with either BRAFi+MEKi or MEKi monotherapy (RR of 56%, Figure 3A). In patients with Class 3 BRAF mutant tumors, the highest response rate was observed with BRAFi+MEKi (RR 27%), whereas either MEKi monotherapy or BRAFi monotherapy were associated with the lowest response rates (RR of 9%, Figure 3A). In multivariable analysis, BRAF Class 2 (aOR 5.836, P=0.001), MEKi (aOR 9.734, P=0.001) and BRAFi+MEKi (aOR 10.947, P<0.001) were independently associated with higher odds of response (Table 2). We explored whether BRAF codon or type of mutation (fusion, internal deletion) were associated with RR, but no apparent trends emerged (Supplemental Figure S6).

**Figure 3:**
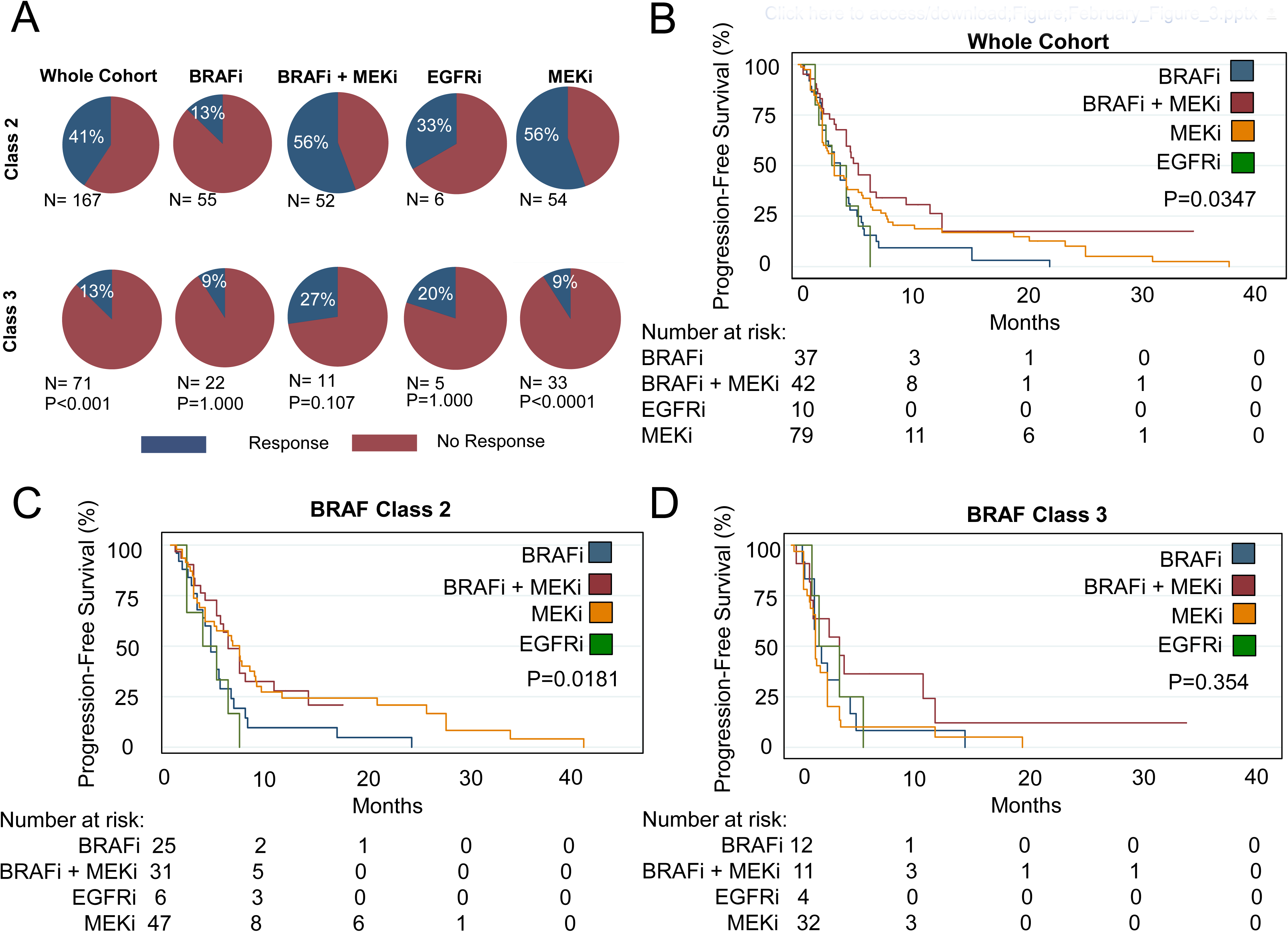
Relationship between BRAF Class and treatment type in the context of MAPK targeted therapy. A) Response rates to MAPK targeted therapy according to BRAF Class and treatment type. P-values calculated with Fischer’s Exact Test. B) Progression-free survival according to treatment type in the entire cohort, and when analyses are restricted to Class 2 (C) and Class 3 (D) BRAF mutant tumors. P-values calculated with Log-Rank test.

In the whole cohort, patients treated with BRAFi + MEKi experienced the longest PFS (mPFS 5.0 months) and those treated with EGFRi experienced the shortest PFS (mPFS 2.8 months, P=0.0347, Figure 3B). In Class 2 mutant tumors, BRAFi + MEKi (mPFS 5.0 months) or MEKi alone (mPFS 6.0 months) were associated with longer PFS compared to BRAFi (mPFS 3.5 months) or EGFRi (mPFS 2.8 months; P=0.0181; Figure 3C). However, in Class 3 mutant tumors, no specific treatment regimen was associated with significantly improved PFS (Figure 3D). In multivariable analysis, BRAFi + MEKi (HR 0.462, 95% CI: 0.27-0.80; P=0.006) and MEKi (HR: 0.588, 0.359-0.96595% CI: 0.36-0.97; P=0.036) were independently associated with longer PFS (Table 3), as was Class 2 BRAF mutational status (HR 0.544, 95% CI: 0.38-0.79; P=0.001). We did not observe a significant association between treatment type and improved outcomes within any of the tumor types analyzed (Table 3, Supplemental Figure S7).

### Depth of response of Class 2 and 3 BRAF mutant tumors to MAPK inhibition is associated with progression-free survival

To better characterize the degree of clinical benefit achieved by patients who responded to MAPKi, we assessed PFS according to response type. Patients who achieved a complete response experienced longer PFS (mPFS 12 months) than patients with partial response (mPFS 6 months), stable disease (mPFS 4.2 months) or progressive disease as best response (mPFS 1.8 months) (Supplemental Figure S8A; P<0.0001). Patients who experienced PFS >12 months demonstrated a greater depth of tumor regression response than patients with responses lasting less than 12 months (Supplemental Figure 8B; P=0.0082). Amongst responders with available data (n=23), we observed a significant correlation between increased depth of response (% tumor regression of target lesions), and longer PFS (Supplemental Figure S8C and D; R^2^=0.2153, P=0.0257).

### Quality Assessment

The majority of the patients included in this analysis were reported in retrospective studies. These retrospective studies may be more subject to bias than prospective studies. Indeed, we observed an increased RR amongst patients reported in retrospective versus prospective studies (42% vs. 13%, P= 0.005; Table 2). To better characterize the risk of bias and its impact on our results, we performed a quality assessment of all studies included in the meta-analysis using a validated 5-point scale (Supplemental Figure S2). We analysed whether risk of bias amongst the studies was associated with treatment response. There was a statistically significant difference in response rate (44% vs. 21%, P<0.001) between patients derived from studies with high risk of bias (score 0-3, n=117) compared with those with low/moderate risk of bias (score 4-5, n=121) (Supplemental Figure S9A); however, risk of bias was not associated with differences in progression-free survival (Supplemental Figure S9B). Amongst studies with low/moderate risk of bias, there was a trend toward response rate being higher amongst patients with Class 2 BRAF mutations (27% vs. 13%) but this difference was not statistically significant (P=0.07; Supplemental Figure S10A). However, the observation that patients with Class 2 BRAF mutations experience longer PFS than patients with Class 3 BRAF mutations was observed in patients from studies with both high and low risk of bias (P=0.0282 and P=0.0194, respectively; Supplemental Figure S10B,C).

## Discussion

By performing a systematic review and meta-analysis of individual patient level data, we have assembled the largest clinical cohort of patients with BRAF non-V600 mutant tumors with associated treatment response to date. This has allowed us to perform comprehensive analyses of characteristics associated with response to MAPK TT in this patient population. The results described herein highlight the importance of testing for the presence of targetable non-V600 BRAF mutations in patients with many types of advanced cancer. These data will be informative for molecular tumor boards and can be used to motivate the design of new clinical trials for patients with non-V600 BRAF mutations.

Like other oncogene driven tumors [33], we observed a strong association between depth of response to oncogene directed therapy and duration of clinical benefit. This finding provides further evidence that Class 2 and 3 BRAF mutations represent key driver oncogenes in these tumors. We found that Class 2 BRAF mutant tumors respond to MAPK targeted therapy more favourably than Class 3 mutants. This finding validates preclinical studies demonstrating that Class 2 BRAF mutant tumors may benefit from therapies that target downstream of mutant RAS while Class 3 mutant tumors require treatment upstream with receptor tyrosine kinase inhibitors [21, 24, 34, 35]. However, there is mounting evidence that Class 2 and Class 3 BRAF mutations can also be important drivers of resistance to EGFRi in colorectal cancer patients [35–37].

In this study, the response rate to MEKi monotherapy or BRAFi + MEKi was 38% and 51%, respectively. This compares favourably to published reports of MEKi monotherapy in RAS mutant lung cancer [38] and melanoma [39], but these comparisons are limited by our analysis of retrospective data and selection bias in case reports and series’. Two previous prospective trials examined the efficacy of MEKi monotherapy (trametinib) for BRAF non-V600 mutant tumors.

The NCI-MATCH (EAY131) study included patients with all primary tumor types and demonstrated a 3% RR [29]. Meanwhile, Nebhan *et al.* included only melanomas with non-V600 BRAF mutations, and observed a 33% RR (3/9) [28].

Despite the fact that the response rates may be higher than expected in this study due to the inclusion of retrospective data, we found no difference in PFS according to whether the data was derived from retrospective vs. prospective studies or high vs. low risk of bias studies. Furthermore, we observed that, in Class 2 BRAF mutant tumors, BRAFi+MEKi or MEKi monotherapy were associated with longer PFS. This provides further evidence that a subset of patients with Class 2 BRAF mutations will derive therapeutic benefit from these treatment regimens. The degree of benefit, in terms of both outcomes and tolerability, conferred by the addition of BRAF inhibition to MEK inhibitors requires further study in prospective trials. Indeed, two on-going clinical trials are currently investigating the efficacy of binimetinib and encorafenib for the treatment of tumors with non-V600 BRAF mutations (NCT03839342, NCT03843775) [40].

In Class 3 BRAF mutant tumors, EGFRi-containing regimens have already been demonstrated to elicit high response rates, particularly when combined with chemotherapy in the context of colorectal cancer [35]. Given that Class 3 mutations may exhibit a degree of additional sensitivity with additional BRAF and / or MEK inhibition, it remains possible that triple therapy regimens, such as the cetuximab, encorafenib and binimetinib combination that proved effective in the BEACON trial for BRAF V600E mutant colorectal cancer may also be beneficial for patients with Class 3 BRAF mutations [34]. Currently, the BIG BANG trial is investigating the efficacy of this combination in colorectal cancer patients with non-V600 BRAF mutations, results that will complement the existing BEACON data [41, 42]. Despite this possibility, it is clear from this dataset that patients with Class 3 BRAF mutant tumors have only modest potential for clinical benefit when treated with currently available standard MAPK targeted therapies. Thus, more research and development of novel therapeutic approaches is urgently needed – particularly for Class 3 BRAF mutations.

We observed a trend towards an association with decreased responsiveness to MAPK targeted therapy in tumors with co-occurring RAS mutations. This observation is perhaps not surprising; RAS mutations are well documented drivers of resistance to EGFR inhibitors in colorectal cancer, and the development of *de novo* RAS mutations has been reported to be a key mechanism of acquired resistance to BRAF +/- MEK inhibitors in BRAF V600 mutant melanoma [43, 44]. Moreover, mutant RAS is capable of activating the PI3K-Akt pathway in addition to the MAPK pathway, potentially promoting resistance to MAPK directed therapy. While these data are only hypothesis-generating, we believe that it will be important in future clinical trials enrolling patients with non-V600 mutations to also report RAS mutation status, as this will help determine if these represent a subset of patients who are unlikely to benefit from currently available MAPK targeted therapies. Recently, a number of drugs targeting KRAS G12C have demonstrated clinical activity, and sotorasib has received FDA approval for the treatment of KRAS G12C mutant non-small cell lung cancer [45]. Several on-going studies are investigating combination therapies for KRAS G12C mutant tumors. Therefore, the possibility of directly targeting KRAS G12C in combination with direct BRAF inhibition in tumors with co-occurring non-V600 BRAF and KRAS mutations is an intriguing therapeutic possibility to overcome RAS-mediated resistance. However, in our study, none of the 14 patients with co-occurring RAS mutations had KRAS G12C mutations, suggesting limited applicability of such a strategy for tumors with non-V600 BRAF mutations.

There are several limitations of this study that are worthy of discussion. First, many patients identified and included in our study are derived from low quality case reports and case series, or small cohorts of patients included in prospective studies. These limitations are exemplified in our comparison of outcomes in patients from retrospective vs. prospective, and low vs. high quality studies. The majority of the patients included in this analysis were from retrospective studies, which reported higher response rates than prospective studies, and were subject to additional bias. As such, the response rates we report in this study likely over-represent the true response rates that would be observed in prospective trials and real-world settings. Interestingly however, progression free survival was not significantly different in retrospective vs. prospective studies, studies that didn’t use RECIST vs. those that did, and studies that were at high risk of bias vs. low risk of bias, suggesting that PFS may be a more reliable, and clinically meaningful metric.

The rarity and variable oncogenic capacity of each individual non-V600 BRAF mutation remains a challenge for drug developers and may complicate interpretation of results, even from future prospective trials. To facilitate effective drug development targeted against these important driver mutations, it will be critical for the oncology community to collaborate in multi-center trials and share data regarding patient responses, tumor types and co-mutation status whenever possible. It is important to note that when examined separately, patients with Class 2 BRAF mutations included in prospective studies or whose response was established with RECIST criteria still demonstrated statistically significant superior PFS compared to those with Class 3 mutations.

Another important limitation of the study is that our analyses are largely based on patients receiving earlier generations of targeted therapies, such as vemurafenib. Emerging preclinical data suggests that alternative BRAF inhibitors such as dabrafenib and encorafenib – both of which can effectively inhibit BRAF dimers to a greater extent than vemurafenib [20] - as well as ‘next-generation’ BRAF dimer inhibitors and pan-RAF inhibitors - which inhibit both BRAF and CRAF - hold substantial promise for non-V600 BRAF mutant tumors [24, 46–48]. In these preclinical studies, differences exist between the efficacy of various MAPK targeted therapies of the same class, but we are underpowered to comment on these differences within this clinical dataset [24, 49]. Finally, our study is limited by the incompleteness of available data in the published literature. Important considerations, such as data on overall survival, performance status, assessment of tumor burden and line of therapy may be important confounders to our results. However, due to insufficient reporting of these parameters in the included publications, they could not be included in our analyses.

Taken together, the existing clinical literature confirms many of the predictions presented by the preclinical studies published over the past two decades with respect to differences between Class 2 and Class 3 BRAF mutants and establishes new hypotheses worthy of further investigation. It is becoming apparent that currently available MAPK targeted therapies have demonstrated clinical activity in a subset of tumors with non-V600 BRAF mutations – especially those with Class 2 BRAF mutations. However, to date, these MAPK-directed therapies appear to be associated with lower response rates than has been observed in patients with BRAF V600 mutant tumors [1, 6, 8, 14, 50, 51]. The efficacy of MAPK inhibitors can be also influenced by tumor type and potentially by co-occuring mutations. Notably, MAPK inhibitor responses are lower in BRAF V600 mutant colorectal cancers than in other BRAF V600 mutated tumor types [1]. As such, more work is needed to better understand the molecular and genomic contexts in which non-V600 BRAF mutant driver oncogenes exist. This could lead to better insight into the molecular mechanisms of intrinsic and acquired resistance to MAPK inhibitors in these tumors. This patient population is heterogeneous and future studies may yield more benefit if therapeutic approaches are tailored according to BRAF Class and primary tumor type. These strategies may include BRAF or pan-RAF inhibitors plus MEK or ERK inhibition for Class 2 mutants and EGFR inhibition (+/- BRAF/pan-RAF/MEK/ERK inhibition) for Class 3 mutants, and BRAF non-V600 mutated colorectal cancers [52]. To date, prospective studies with targeted monotherapies have yielded modest response rates [28, 29]. These data clearly suggest that future clinical trials aimed at developing drugs to target tumors with non-V600 BRAF mutations should incorporate combination therapy strategies.

## Data Availability

All data produced in the present study are available upon reasonable request to the authors

## Acknowledgements

MD and YW acknowledge support from Vanier Canada Graduate Scholarships. GZ and AANR acknowledges support from a Fonds de Recherche du Québec – Santé (FRQS) Clinical Research Scholar award. AANR is a recipient of a Conquer Cancer Foundation of ASCO Young Investigator Award.

---

**Supplemental Figure S1:**
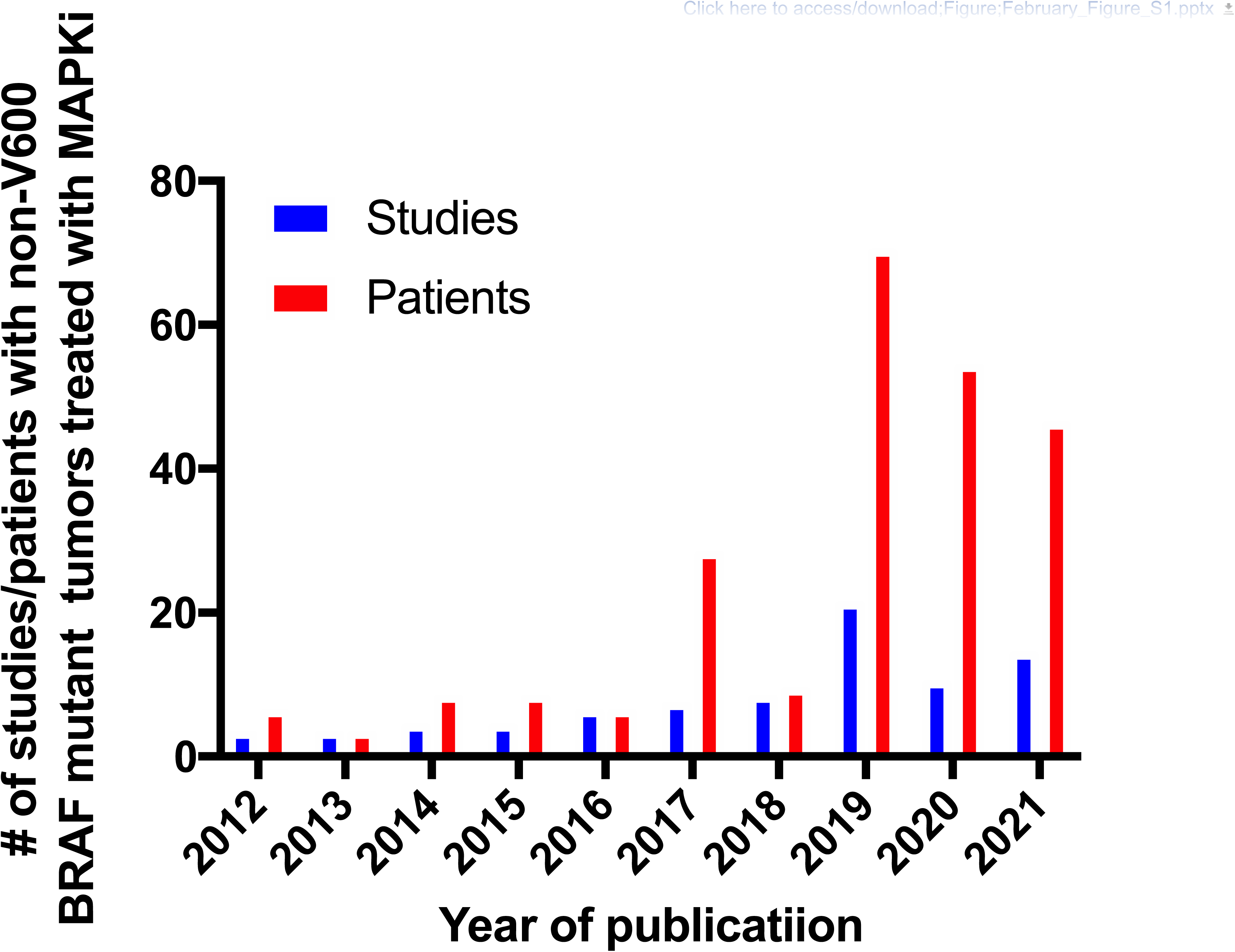
Description of studies and patients included in the meta-analysis by year of publication.

**Supplemental Figure S2.**
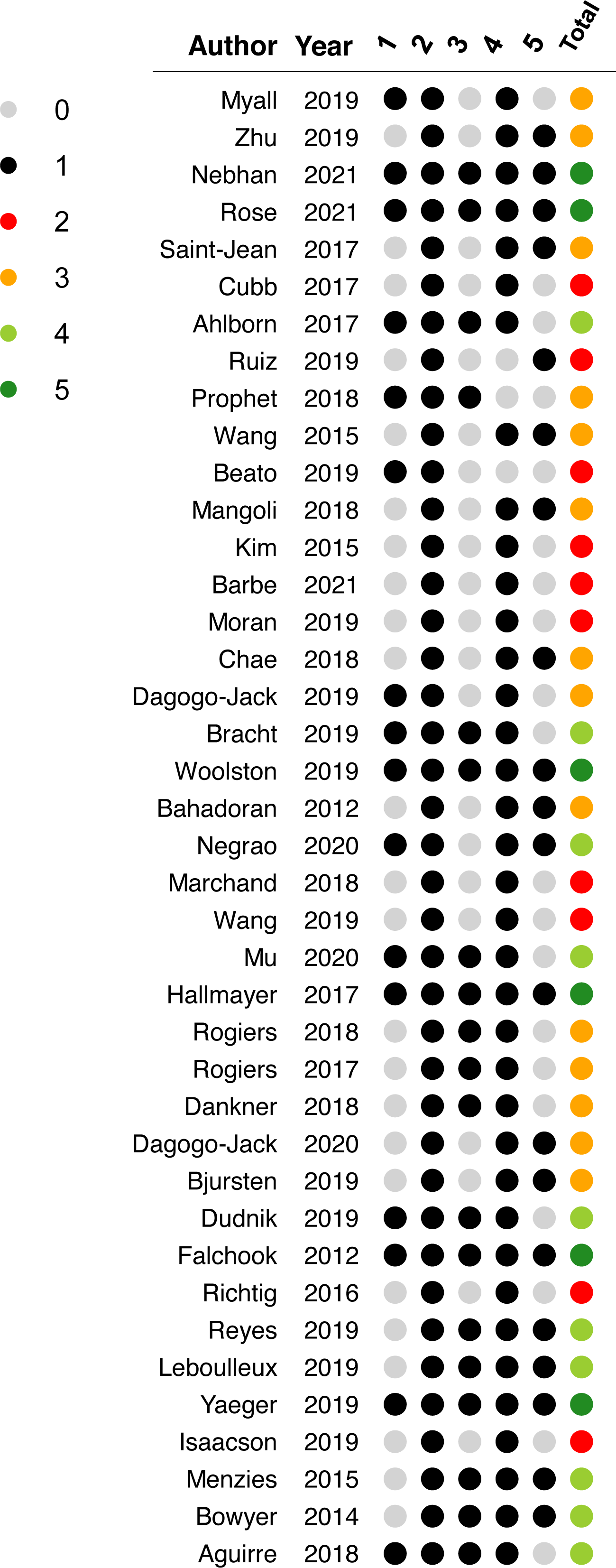
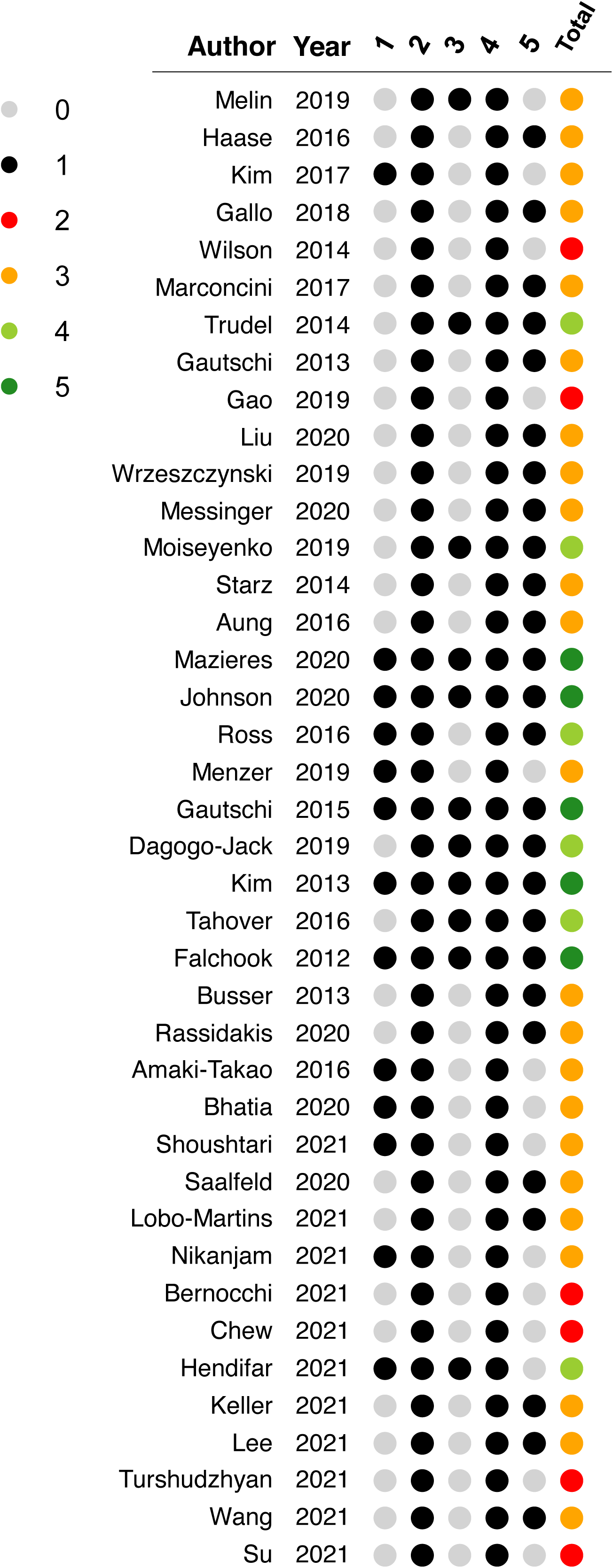
Risk of bias assessment of the individual studies included in the systematic review. The 5-point score is adapted from the Newcastle-Ottawa score: 1) Selection – Did the patients represent all/consecutive patients with non-V600 BRAF mutations from the medical center? 2) Ascertainment (Diagnosis) – Was the diagnosis correctly made with pathology-proven cancer and next-generation sequencing assay to confirm BRAF mutation? 3) Ascertainment (Outcome) – Was treatment response adequately ascertained using RECIST criteria? 4) Follow-up – Was follow up long enough for treatment responses to be evaluated? 5) Reporting – Is the case described with sufficient details (*e.g.,* drug posology, previous lines of chemotherapy) to allow other investigators to replicate the research or to allow practitioners make inferences related to their own practice? The 5 first columns represent the 5 assessment criteria (black circle = yes, grey circle = no). The total risk of bias score is the last column and the colors indicate the score.

**Supplemental Figure S3:**
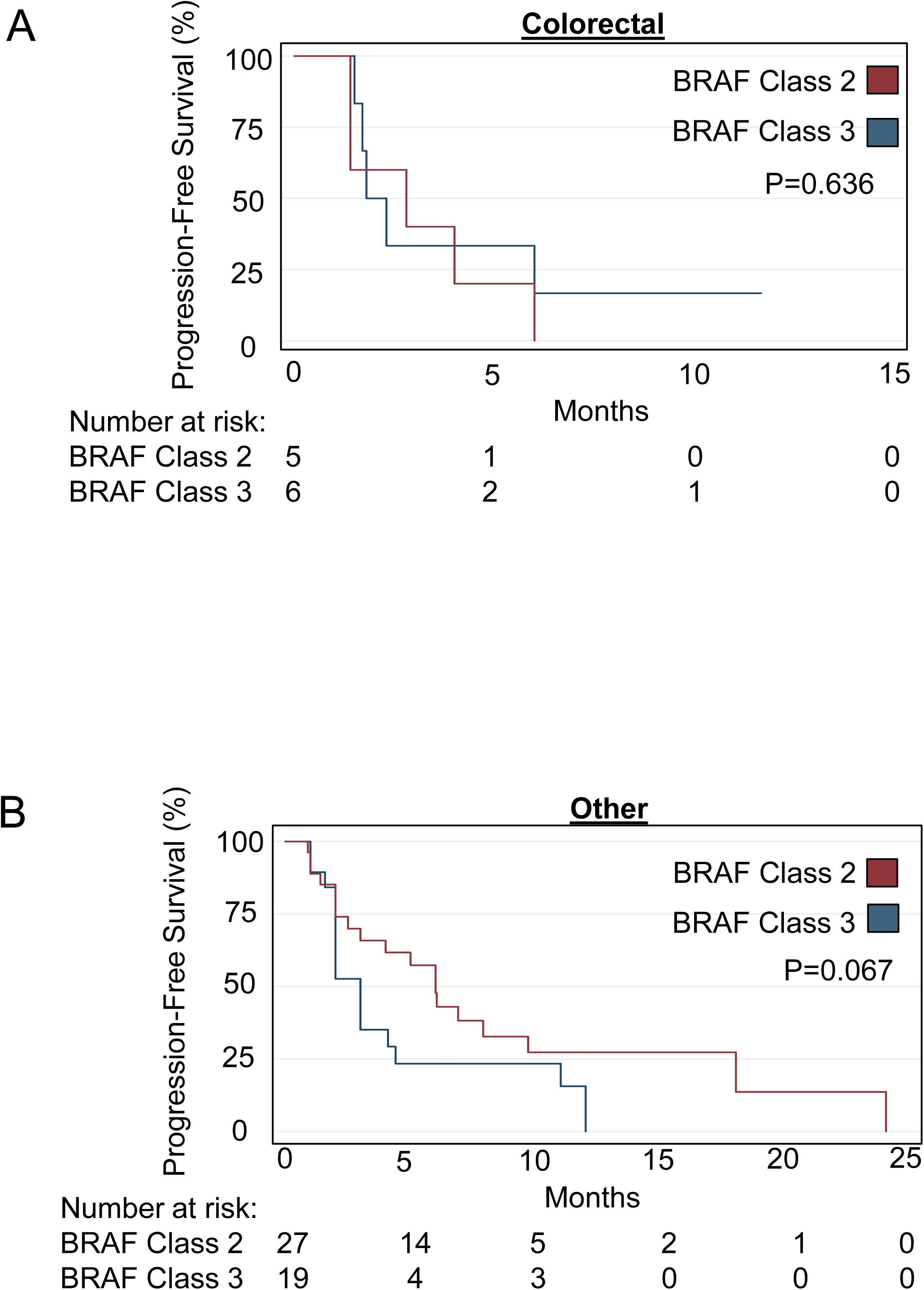
Progression-free survival according to BRAF Class in (A) colorectal and (B) non-melanoma/lung/colorectal ‘other’ primary tumors. P-values calculated with Log-Rank test.

**Supplemental Figure S4:**
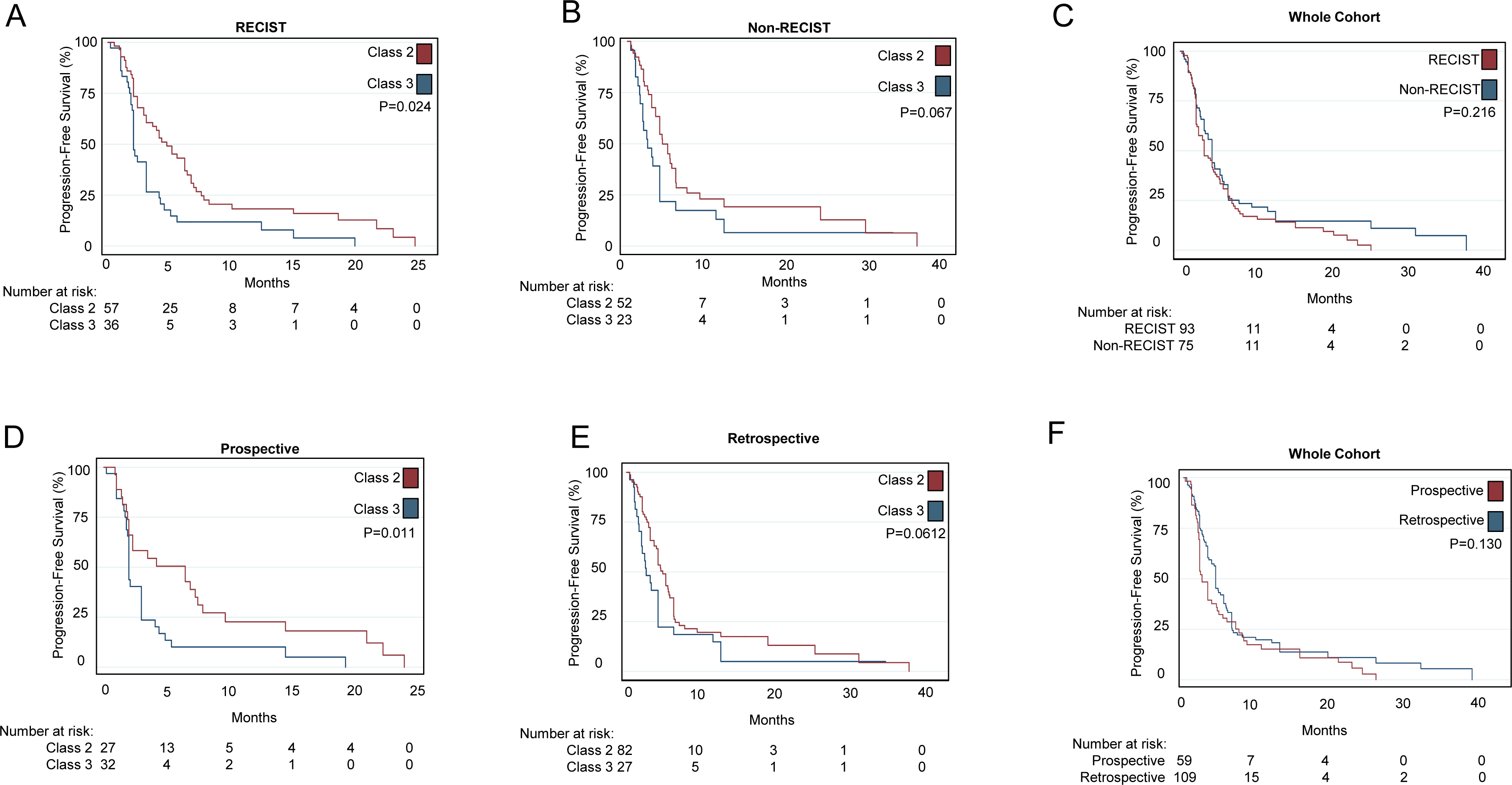
Progression-free survival according to (A, B,C) response criteria type and (D, E, F) study type. P-values calculated with Log-Rank test.

**Supplemental Figure S5:**
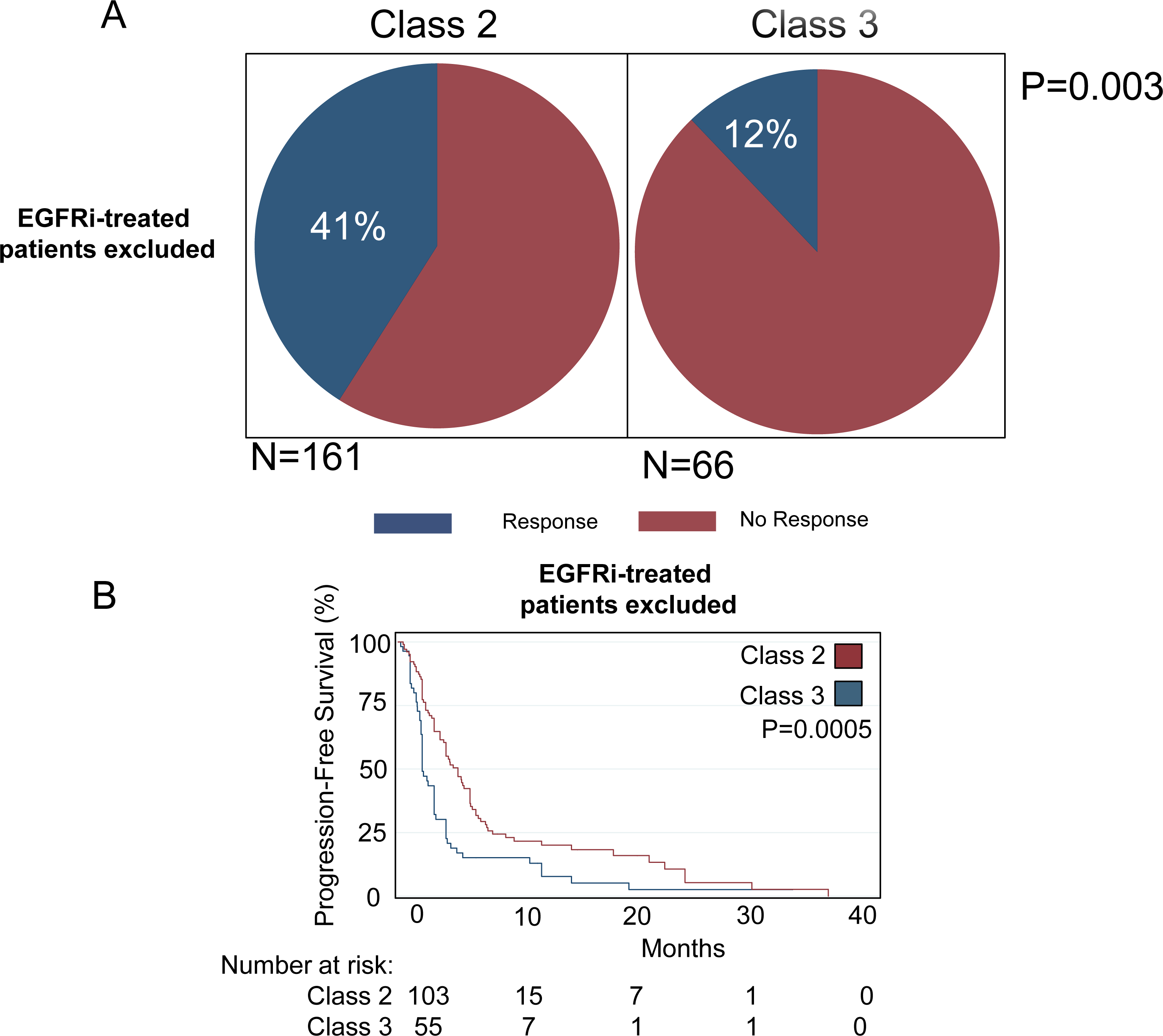
Response rate (A) and progression-free survival (B) between BRAF mutant Classes in MEKi+/-BRAFi treated patients. P-values calculated with Fischer’s Exact test (A) and Log-Rank test (B).

**Supplemental Figure S6:**
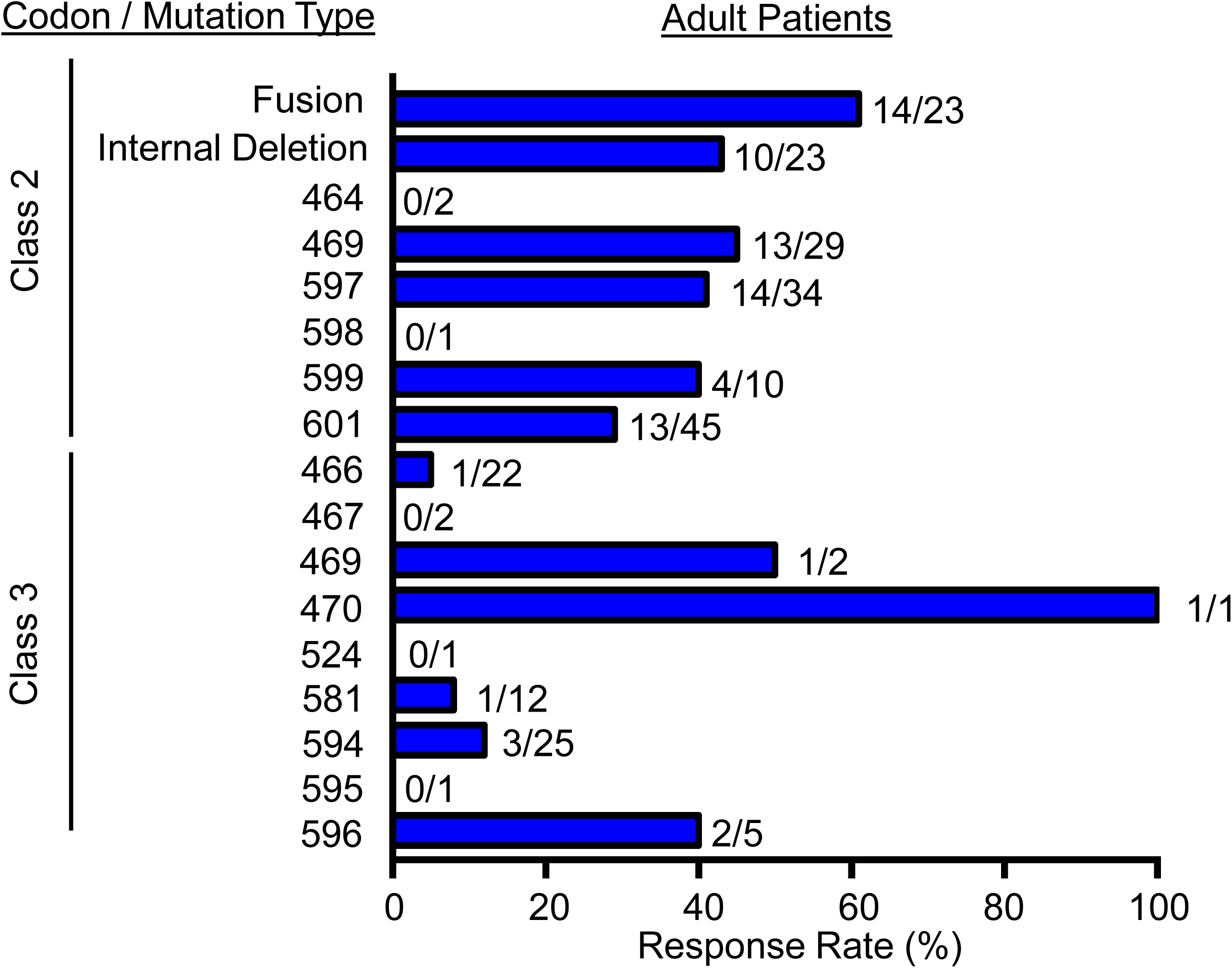
Response rate in the study cohort by individual codon mutated or other type of mutation (fusion, internal deletion) detected.

**Supplemental Figure S7:**
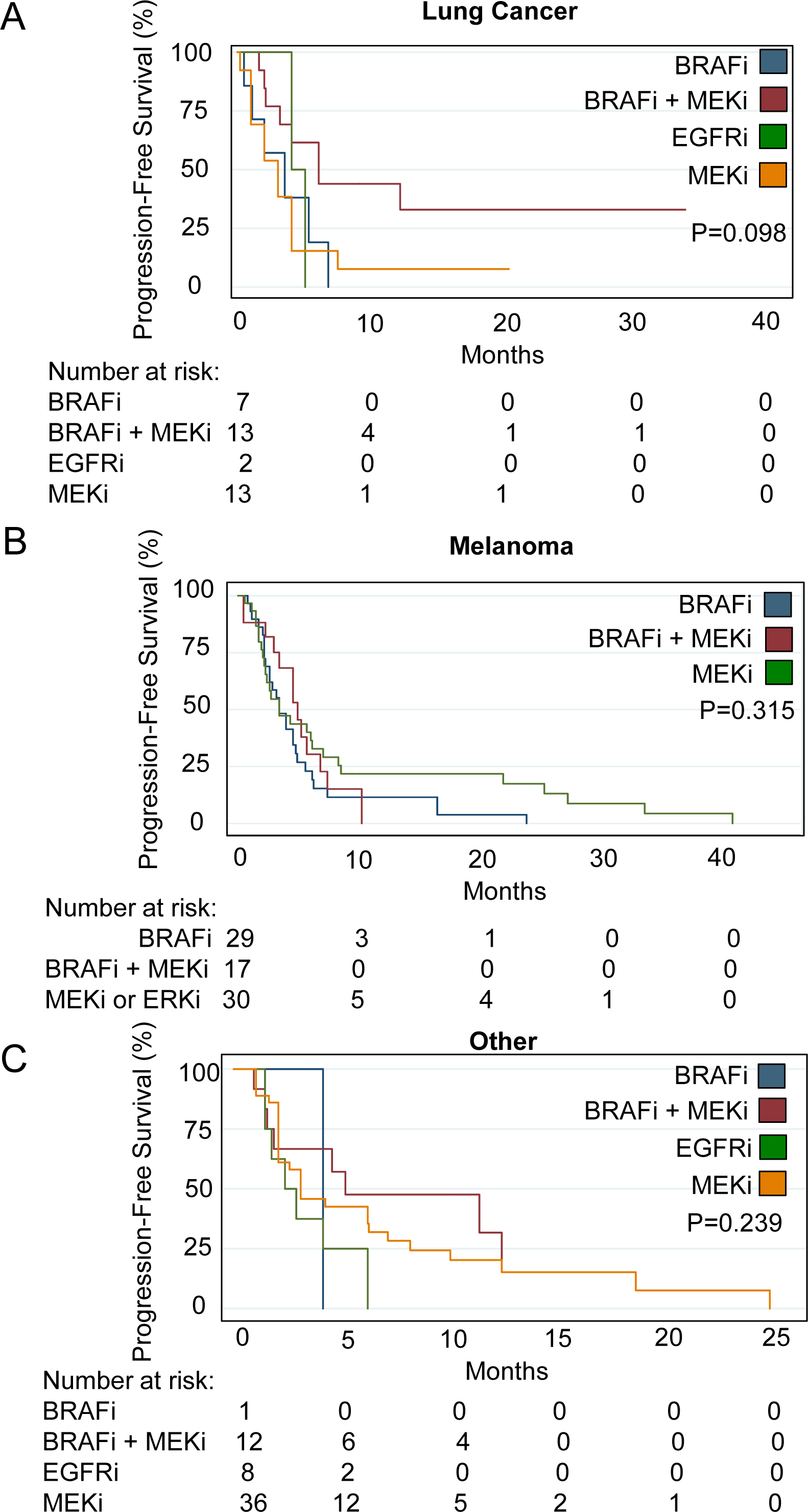
Progression-free survival stratified according to primary tumor type. (A) lung cancer primary, (B) melanoma primary and (C) non-lung non-melanoma ‘other’ primary tumors. P-values calculated with Log-Rank test.

**Supplemental Figure S8:**
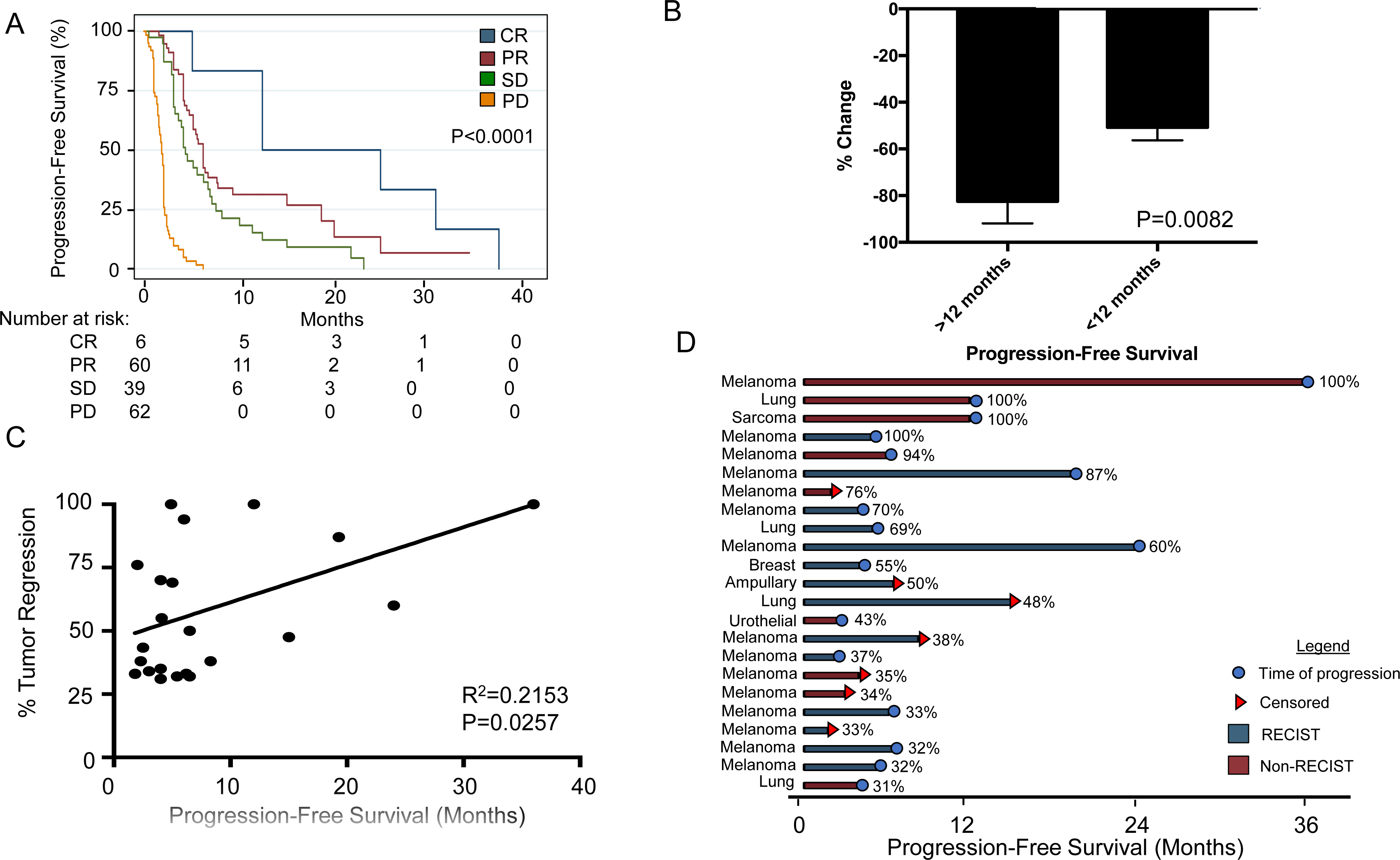
Relationship between depth and duration of response. A) Kaplan-Meier curve demonstrating progression-free survival plotted by type of response: complete response (CR), partial response (PR), stable disease (SD) and progressive disease (PD). P-value calculated with Log-Rank test. B) Patients with short (<12 months) or long (>12 months) responses plotted by depth of response (% tumor size change). P-value calculated with 2-tailed Student’s T-Test. C) Correlation between % tumor regression and progression-free survival among patients who responded to therapy (PR, CR) and who had % regression data available. P-value calculated by linear regression. D) Swimmer’s plot of all patients who responded to therapy (CR, PR) and who had available % regression data available. Primary tumor type is described on the left, color of the bar represents whether the response was determined with RECIST criteria or not, the circle or triangle at the end of the bar represents progression or censoring, and the percentage at the far right of the bar refers to the depth of response (% tumor regression).

**Supplemental Figure S9:**
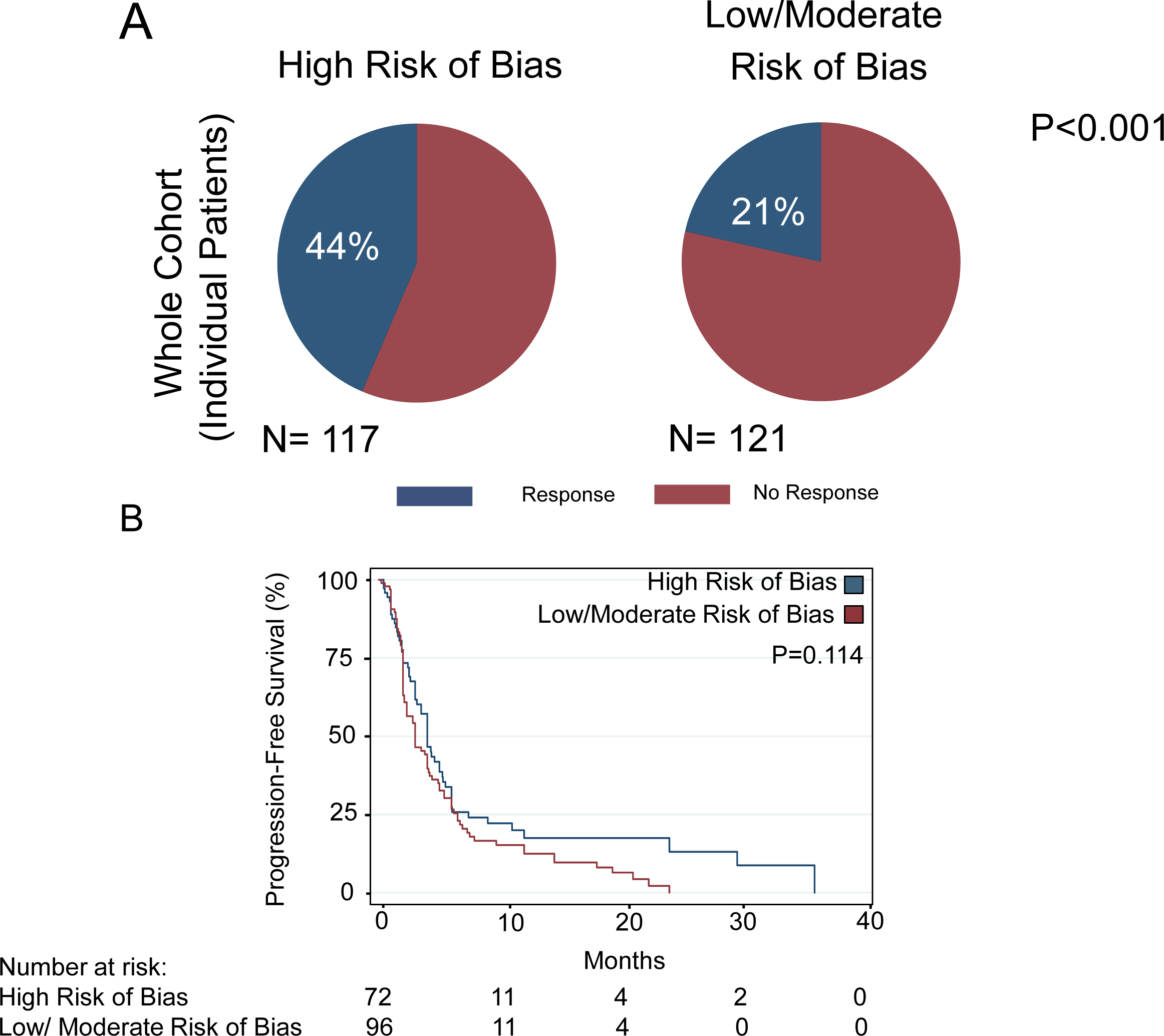
Patients from studies with high vs. low/moderate risk of bias by (A) response rates and (B) progression-free survival. P-value calculated with Fischer’s Exact test (A) and Log-Rank test (B).

**Supplemental Figure S10:**
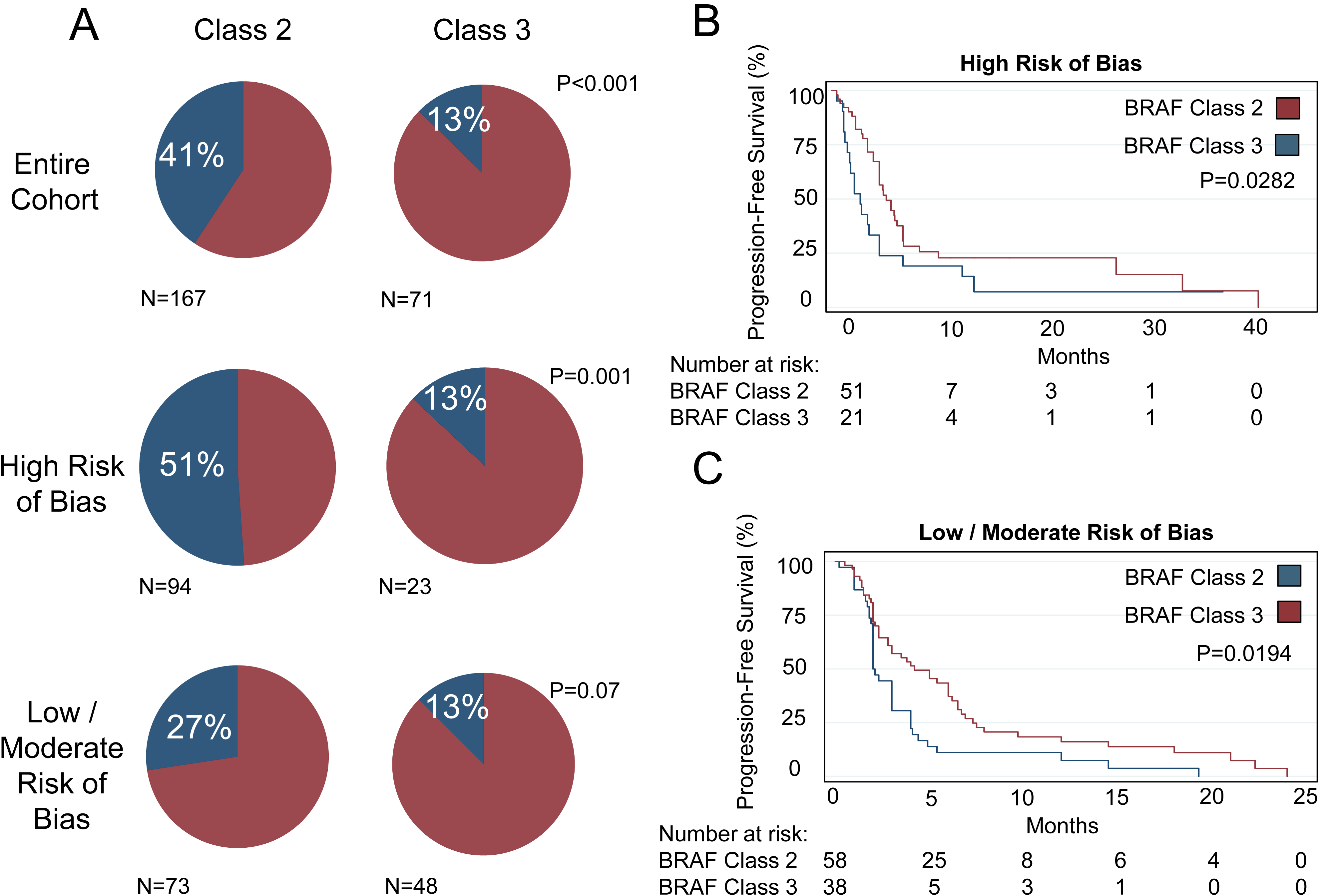
A) Response rate stratified by BRAF Class in the entire cohort (top), studies with high risk of bias (middle) and studies with low/moderate risk of bias (bottom). B) Kaplan-Meier curves of progression-free survival by BRAF Class in patients from studies with high risk of bias and C) low/moderate risk of bias. P-value calculated with (A) Fischer’s Exact test, (B,C) Log-Rank test.

**Supplemental Table S1.** MAPK targeted therapy regimens used for all patients in the study. FDA-approved monomeric BRAFi includes vemurafenib, dabrafenib or encorafenib, and FDA-approved MEKi includes cobimetinib, trametinib, binimetinib or selumetinib.

|  |  |  |
| --- | --- | --- |
| **MAPKi Class** | **MAPKi Regimen** | **# of Patients** |
| **BRAFi** | Vemurafenib | 51 |
|  | Dabrafenib | 9 |
|  | Unknown FDA-approved monomeric BRAFi | 17 |
| **MEKi** | Cobimetinib | 2 |
|  | Trametinib | 80 |
|  | Selumetinib | 2 |
|  | Binimetinib | 0 |
|  | Unknown FDA-approved MEKi | 3 |
| **BRAFi + MEKi** | Vemurafenib + Cobimetinib | 3 |
|  | Dabrafenib + Trametinib | 30 |
|  | Encorafenib + Binimetinib | 8 |
|  | Unknown FDA-approved BRAFi + MEKi | 22 |
| **EGFRi** | Cetuximab | 4 |
|  | Panitumumab | 4 |
|  | Osimertinib | 1 |
|  | Binimetinib + Panitumumab | 1 |
|  | Osimertinib + Trametinib | 1 |

**Supplemental Table S2.** List of Class 2 and Class 3 mutations included in the study

| BRAF Class | Mutation |  |
| --- | --- | --- |
| 2 | L597V/S/R/Q/P/K | L525R |
|  | K601E/N | L485W/F |
|  | A598V/T599insV | V600_K601del |
|  | T599I/dup/V600insT | V600_K601D/E/N |
|  | G464V/E | V600_K602delinsDT |
|  | N486_P490del | V600_W604delinsDQTDG |
|  | G469V/S/R/L/A/T170delinsAK | BRAF Fusions |
| 3 | D594N/G/F | F595L |
|  | G466E/V/A | T470R |
|  | N581S/T/I | Q524L |
|  | G596V/R |  |
|  | G469E |  |
|  | S467L |  |

**Supplemental Table S3.** Fusion partners of BRAF fusions identified in the study

| **Fusion Partner** | **Number of Patients** |
| --- | --- |
| AGAP3 | 1 |
| AGK | 1 |
| KHDRBS2 | 0 |
| KIAA1549 | 3 |
| KMT2E | 1 |
| MZT1 | 1 |
| NRF1 | 1 |
| PDE10A | 0 |
| PICALM | 1 |
| PPFIBP2 | 1 |
| PWWP2A | 1 |
| SLC45A3 | 2 |
| SND1 | 2 |
| TRIM24 | 1 |
| ZKSCAN1 | 1 |
| ZNF767 | 1 |
| LIMD1 | 1 |
| SKAP2 | 1 |
| Unknown Fusion Partner | 0 |

Appendix 1: Detailed search strategy.

Appendix 2: List of references of studies used to extract data for the systematic review and meta-analysis.

