## Appendix 1 for "Clinical activity of MAPK targeted therapies in patients with non-V600 BRAF mutant tumors"

**Ovid MEDLINE(R) ALL 2010** **to September 13, 2021**

| **#** | **Searches** | **Results** | **Type** |
| --- | --- | --- | --- |
| 1 | Proto-Oncogene Proteins B-raf/ | 9762 | Advanced |
| 2 | b-raf*.mp,kw. | 10802 | Advanced |
| 3 | (braf* adj10 alternative*).mp,kw. | 156 | Advanced |
| 4 | (braf* adj10 atypical*).mp,kw. | 70 | Advanced |
| 5 | (braf* adj10 class*2).mp,kw. | 201 | Advanced |
| 6 | (braf* adj10 deletion?).mp,kw. | 175 | Advanced |
| 7 | (braf* adj10 fusion?).mp,kw. | 601 | Advanced |
| 8 | (braf* adj10 muta?t*).mp,kw. | 12534 | Advanced |
| 9 | (braf* adj10 non-canonical?).mp,kw. | 7 | Advanced |
| 10 | (braf* adj10 noncanonical?).mp,kw. | 5 | Advanced |
| 11 | V600*.mp,kw. | 5194 | Advanced |
| 12 | brafv600*.mp,kw. | 1914 | Advanced |
| 13 | b-rafv600*.mp,kw. | 73 | Advanced |
| 14 | V-600*.mp,kw. | 78 | Advanced |
| 15 | brafv-600*.mp,kw. | 10 | Advanced |
| 16 | b-rafv-600*.mp,kw. | 1 | Advanced |
| 17 | nonV600*.mp,kw. | 2 | Advanced |
| 18 | brafnonv600*.mp,kw. | 2 | Advanced |
| 19 | b-rafnonv600*.mp,kw. | 0 | Advanced |
| 20 | nonV-600*.mp,kw. | 0 | Advanced |
| 21 | brafnonv-600*.mp,kw. | 0 | Advanced |
| 22 | b-rafnonv-600*.mp,kw. | 0 | Advanced |
| 23 | A598*.mp,kw. | 11 | Advanced |
| 24 | brafa598*.mp,kw. | 2 | Advanced |
| 25 | b-rafa598*.mp,kw. | 0 | Advanced |
| 26 | A727*.mp,kw. | 11 | Advanced |
| 27 | brafa727*.mp,kw. | 0 | Advanced |
| 28 | b-rafa727*.mp,kw. | 0 | Advanced |
| 29 | D594*.mp,kw. | 35 | Advanced |
| 30 | brafd594*.mp,kw. | 3 | Advanced |
| 31 | b-rafd594*.mp,kw. | 1 | Advanced |
| 32 | E586*.mp,kw. | 8 | Advanced |
| 33 | brafe586*.mp,kw. | 0 | Advanced |
| 34 | b-rafe586*.mp,kw. | 0 | Advanced |
| 35 | F595*.mp,kw. | 19 | Advanced |
| 36 | braff595*.mp,kw. | 0 | Advanced |
| 37 | b-raff595*.mp,kw. | 0 | Advanced |
| 38 | G464*.mp,kw. | 43 | Advanced |
| 39 | brafg464*.mp,kw. | 0 | Advanced |
| 40 | b-rafg464*.mp,kw. | 0 | Advanced |
| 41 | G466*.mp,kw. | 36 | Advanced |
| 42 | brafg466*.mp,kw. | 2 | Advanced |
| 43 | b-rafg466*.mp,kw. | 0 | Advanced |
| 44 | G469*.mp,kw. | 51 | Advanced |
| 45 | brafg469*.mp,kw. | 4 | Advanced |
| 46 | b-rafg469*.mp,kw. | 0 | Advanced |
| 47 | G596*.mp,kw. | 18 | Advanced |
| 48 | brafg596*.mp,kw. | 0 | Advanced |
| 49 | b-rafg596*.mp,kw. | 0 | Advanced |
| 50 | I463*.mp,kw. | 9 | Advanced |
| 51 | brafi463*.mp,kw. | 0 | Advanced |
| 52 | b-rafi463*.mp,kw. | 0 | Advanced |
| 53 | K483*.mp,kw. | 11 | Advanced |
| 54 | brafk483*.mp,kw. | 0 | Advanced |
| 55 | b-rafk483*.mp,kw. | 0 | Advanced |
| 56 | K601*.mp,kw. | 100 | Advanced |
| 57 | brafk601*.mp,kw. | 9 | Advanced |
| 58 | b-rafk601*.mp,kw. | 1 | Advanced |
| 59 | L597*.mp,kw. | 28 | Advanced |
| 60 | brafl597*.mp,kw. | 2 | Advanced |
| 61 | b-rafl597*.mp,kw. | 0 | Advanced |
| 62 | N581*.mp,kw. | 78 | Advanced |
| 63 | brafn581*.mp,kw. | 0 | Advanced |
| 64 | b-rafn581*.mp,kw. | 0 | Advanced |
| 65 | R462*.mp,kw. | 46 | Advanced |
| 66 | brafr462*.mp,kw. | 0 | Advanced |
| 67 | b-rafr462*.mp,kw. | 0 | Advanced |
| 68 | S467*.mp,kw. | 20 | Advanced |
| 69 | brafs467*.mp,kw. | 0 | Advanced |
| 70 | b-rafs467*.mp,kw. | 0 | Advanced |
| 71 | T599*.mp,kw. | 36 | Advanced |
| 72 | braft599*.mp,kw. | 1 | Advanced |
| 73 | b-raft599*.mp,kw. | 0 | Advanced |
| 74 | or/1-73 | 17488 | Advanced |
| 75 | Gefitinib/ | 4695 | Advanced |
| 76 | Cetuximab/ | 4833 | Advanced |
| 77 | exp Trastuzumab/ | 7608 | Advanced |
| 78 | Vemurafenib/ | 1447 | Advanced |
| 79 | (map* adj3 inhibitor?).mp,kw. | 9781 | Advanced |
| 80 | (mek* adj3 inhibitor?).mp,kw. | 9405 | Advanced |
| 81 | (erk* adj3 inhibitor?).mp,kw. | 7836 | Advanced |
| 82 | (raf adj3 inhibitor?).mp,kw. | 1708 | Advanced |
| 83 | (braf* adj3 inhibitor?).mp,kw. | 3349 | Advanced |
| 84 | (b-raf* adj3 inhibitor?).mp,kw. | 326 | Advanced |
| 85 | mapk?.mp,kw. | 71707 | Advanced |
| 86 | mek?.mp,kw. | 24798 | Advanced |
| 87 | erk?.mp,kw. | 82378 | Advanced |
| 88 | map*kinase?.mp,kw. | 1144 | Advanced |
| 89 | mapkkk*.mp,kw. | 663 | Advanced |
| 90 | mapk-erk?.mp,kw. | 4644 | Advanced |
| 91 | mapk*erk*.mp,kw. | 331 | Advanced |
| 92 | mekk?.mp,kw. | 1206 | Advanced |
| 93 | map?k?.mp,kw. | 76641 | Advanced |
| 94 | pan-raf.mp,kw. | 75 | Advanced |
| 95 | panraf.mp,kw. | 3 | Advanced |
| 96 | afatinib*.mp,kw. | 1664 | Advanced |
| 97 | bibw 2992.mp,kw. | 60 | Advanced |
| 98 | bibw2992.mp,kw. | 24 | Advanced |
| 99 | gilotrif*.mp,kw. | 14 | Advanced |
| 100 | giotrif*.mp,kw. | 8 | Advanced |
| 101 | 41ud74l59m.rn. | 792 | Advanced |
| 102 | 439081-18-2.rn. | 0 | Advanced |
| 103 | 850140-72-6.rn. | 0 | Advanced |
| 104 | 850140-73-7.rn. | 0 | Advanced |
| 105 | erlotinib*.mp,kw. | 7300 | Advanced |
| 106 | cp 358774.mp,kw. | 4 | Advanced |
| 107 | cp358774.mp,kw. | 2 | Advanced |
| 108 | nsc 718781.mp,kw. | 1 | Advanced |
| 109 | nsc71878.mp,kw. | 0 | Advanced |
| 110 | osi 774.mp,kw. | 104 | Advanced |
| 111 | osi774.mp,kw. | 1 | Advanced |
| 112 | r 1415.mp,kw. | 1 | Advanced |
| 113 | r1415.mp,kw. | 0 | Advanced |
| 114 | tarceva*.mp,kw. | 329 | Advanced |
| 115 | da87705x9k.rn. | 4112 | Advanced |
| 116 | 183319-69-9.rn. | 0 | Advanced |
| 117 | 183321-74-6.rn. | 0 | Advanced |
| 118 | gefitinib*.mp,kw. | 7693 | Advanced |
| 119 | geftinat*.mp,kw. | 0 | Advanced |
| 120 | iressa*.mp,kw. | 816 | Advanced |
| 121 | zd 1839.mp,kw. | 43 | Advanced |
| 122 | zd1839.mp,kw. | 447 | Advanced |
| 123 | s65743jhbs.rn. | 4695 | Advanced |
| 124 | 184475-35-2.rn. | 0 | Advanced |
| 125 | 184475-55-6.rn. | 0 | Advanced |
| 126 | 184475-56-7.rn. | 0 | Advanced |
| 127 | osimertinib*.mp,kw. | 1603 | Advanced |
| 128 | azd 9291.mp,kw. | 10 | Advanced |
| 129 | azd9291.mp,kw. | 199 | Advanced |
| 130 | tagrisso*.mp,kw. | 26 | Advanced |
| 131 | 1421373-65-0.rn. | 0 | Advanced |
| 132 | 1421373-66-1.rn. | 0 | Advanced |
| 133 | cetuximab*.mp,kw. | 7916 | Advanced |
| 134 | c225.mp,kw. | 494 | Advanced |
| 135 | c 225.mp,kw. | 80 | Advanced |
| 136 | erbitux*.mp,kw. | 351 | Advanced |
| 137 | imcc225.mp,kw. | 0 | Advanced |
| 138 | imcc 225.mp,kw. | 0 | Advanced |
| 139 | pqx0d8j21j.rn. | 4833 | Advanced |
| 140 | 205923-56-4.rn. | 0 | Advanced |
| 141 | trastuzumab*.mp,kw. | 12563 | Advanced |
| 142 | abp 980.mp,kw. | 12 | Advanced |
| 143 | abp980.mp,kw. | 1 | Advanced |
| 144 | aryotrust*.mp,kw. | 1 | Advanced |
| 145 | "bcd 022".mp,kw. | 1 | Advanced |
| 146 | bcd022.mp,kw. | 0 | Advanced |
| 147 | bx 2318.mp,kw. | 0 | Advanced |
| 148 | bx2318.mp,kw. | 0 | Advanced |
| 149 | ct p06.mp,kw. | 0 | Advanced |
| 150 | ctp06.mp,kw. | 0 | Advanced |
| 151 | ct p6.mp,kw. | 15 | Advanced |
| 152 | ctp6.mp,kw. | 1 | Advanced |
| 153 | da 3111.mp,kw. | 0 | Advanced |
| 154 | da3111.mp,kw. | 0 | Advanced |
| 155 | dmb 3111.mp,kw. | 1 | Advanced |
| 156 | dmb3111.mp,kw. | 0 | Advanced |
| 157 | eg 12014.mp,kw. | 0 | Advanced |
| 158 | eg12014.mp,kw. | 0 | Advanced |
| 159 | gb 221.mp,kw. | 0 | Advanced |
| 160 | gb221.mp,kw. | 0 | Advanced |
| 161 | hd 201.mp,kw. | 2 | Advanced |
| 162 | hd201.mp,kw. | 3 | Advanced |
| 163 | herceptin*.mp,kw. | 2026 | Advanced |
| 164 | hermyl 1401o.mp,kw. | 0 | Advanced |
| 165 | hermyl1401o.mp,kw. | 0 | Advanced |
| 166 | herticad*.mp,kw. | 0 | Advanced |
| 167 | hertraz*.mp,kw. | 1 | Advanced |
| 168 | hervelous*.mp,kw. | 0 | Advanced |
| 169 | herzuma*.mp,kw. | 9 | Advanced |
| 170 | "hlx 02".mp,kw. | 0 | Advanced |
| 171 | hlx02.mp,kw. | 3 | Advanced |
| 172 | kanjinti*.mp,kw. | 6 | Advanced |
| 173 | myl 1401o.mp,kw. | 6 | Advanced |
| 174 | myl1401o.mp,kw. | 0 | Advanced |
| 175 | ogivri*.mp,kw. | 7 | Advanced |
| 176 | ons 1050.mp,kw. | 0 | Advanced |
| 177 | ons1050.mp,kw. | 0 | Advanced |
| 178 | ontruzant*.mp,kw. | 8 | Advanced |
| 179 | "pf 05280014".mp,kw. | 10 | Advanced |
| 180 | pf05280014.mp,kw. | 0 | Advanced |
| 181 | pf 5280014.mp,kw. | 0 | Advanced |
| 182 | pf5280014.mp,kw. | 0 | Advanced |
| 183 | r 597.mp,kw. | 15 | Advanced |
| 184 | r597.mp,kw. | 5 | Advanced |
| 185 | rg 597.mp,kw. | 0 | Advanced |
| 186 | rg597.mp,kw. | 0 | Advanced |
| 187 | sb 3.mp,kw. | 205 | Advanced |
| 188 | sb3.mp,kw. | 295 | Advanced |
| 189 | trasturel*.mp,kw. | 2 | Advanced |
| 190 | trazimera*.mp,kw. | 3 | Advanced |
| 191 | "tx 05".mp,kw. | 5 | Advanced |
| 192 | tx05.mp,kw. | 1 | Advanced |
| 193 | ub 921.mp,kw. | 0 | Advanced |
| 194 | ub921.mp,kw. | 0 | Advanced |
| 195 | vivitra*.mp,kw. | 1 | Advanced |
| 196 | zedora*.mp,kw. | 3 | Advanced |
| 197 | zrc 3256.mp,kw. | 0 | Advanced |
| 198 | zrc3256.mp,kw. | 0 | Advanced |
| 199 | p188anx8ck.rn. | 7538 | Advanced |
| 200 | 1446410-98-5.rn. | 0 | Advanced |
| 201 | 180288-69-1.rn. | 0 | Advanced |
| 202 | belvarafenib*.mp,kw. | 3 | Advanced |
| 203 | gdc5573.mp,kw. | 0 | Advanced |
| 204 | gdc-5573.mp,kw. | 0 | Advanced |
| 205 | lxh254.mp,kw. | 6 | Advanced |
| 206 | lxh-254.mp,kw. | 0 | Advanced |
| 207 | hm95573.mp,kw. | 0 | Advanced |
| 208 | hm-95573.mp,kw. | 0 | Advanced |
| 209 | rg6185.mp,kw. | 0 | Advanced |
| 210 | rg-6185.mp,kw. | 0 | Advanced |
| 211 | 1446113-23-0.rn. | 0 | Advanced |
| 212 | binimetinib*.mp,kw. | 242 | Advanced |
| 213 | balimek*.mp,kw. | 0 | Advanced |
| 214 | mek 162.mp,kw. | 11 | Advanced |
| 215 | mek162.mp,kw. | 66 | Advanced |
| 216 | mektovi*.mp,kw. | 6 | Advanced |
| 217 | 1073666-70-2.rn. | 0 | Advanced |
| 218 | 120168-50-5.rn. | 0 | Advanced |
| 219 | 606143-89-9.rn. | 0 | Advanced |
| 220 | cobimetinib*.mp,kw. | 348 | Advanced |
| 221 | cotellic*.mp,kw. | 5 | Advanced |
| 222 | "gdc 0973".mp,kw. | 25 | Advanced |
| 223 | gdc-0973.mp,kw. | 25 | Advanced |
| 224 | rg 7420.mp,kw. | 2 | Advanced |
| 225 | rg7420.mp,kw. | 0 | Advanced |
| 226 | xl 518.mp,kw. | 1 | Advanced |
| 227 | xl518.mp,kw. | 5 | Advanced |
| 228 | 1369665-02-0.rn. | 0 | Advanced |
| 229 | 934660-93-2.rn. | 0 | Advanced |
| 230 | dabrafenib*.mp,kw. | 1388 | Advanced |
| 231 | gsk 2118436?.mp,kw. | 3 | Advanced |
| 232 | gsk2118436?.mp,kw. | 33 | Advanced |
| 233 | tafinlar*.mp,kw. | 16 | Advanced |
| 234 | 1195765-45-7.rn. | 0 | Advanced |
| 235 | 1195768-06-9.rn. | 0 | Advanced |
| 236 | encorafenib*.mp,kw. | 197 | Advanced |
| 237 | braftovi*.mp,kw. | 9 | Advanced |
| 238 | lgx 818.mp,kw. | 3 | Advanced |
| 239 | lgx818.mp,kw. | 19 | Advanced |
| 240 | 1269440-17-6.rn. | 0 | Advanced |
| 241 | trametinib*.mp,kw. | 1570 | Advanced |
| 242 | gsk 1120212?.mp,kw. | 6 | Advanced |
| 243 | gsk1120212?.mp,kw. | 55 | Advanced |
| 244 | jtp 74057.mp,kw. | 6 | Advanced |
| 245 | jtp74057.mp,kw. | 0 | Advanced |
| 246 | mekinist*.mp,kw. | 11 | Advanced |
| 247 | 1187431-43-1.rn. | 0 | Advanced |
| 248 | 871700-17-3.rn. | 0 | Advanced |
| 249 | ulixertinib*.mp,kw. | 24 | Advanced |
| 250 | bvd 523.mp,kw. | 9 | Advanced |
| 251 | bvd523.mp,kw. | 0 | Advanced |
| 252 | hy 15816.mp,kw. | 0 | Advanced |
| 253 | hy15816.mp,kw. | 0 | Advanced |
| 254 | vrt 752271.mp,kw. | 0 | Advanced |
| 255 | vrt752271.mp,kw. | 0 | Advanced |
| 256 | 869886-67-9.rn. | 0 | Advanced |
| 257 | vemurafenib*.mp,kw. | 2583 | Advanced |
| 258 | plx 4032.mp,kw. | 25 | Advanced |
| 259 | plx4032.mp,kw. | 183 | Advanced |
| 260 | r 7204.mp,kw. | 0 | Advanced |
| 261 | r7204.mp,kw. | 0 | Advanced |
| 262 | rg 7204.mp,kw. | 1 | Advanced |
| 263 | rg7204.mp,kw. | 22 | Advanced |
| 264 | ro 5185426.mp,kw. | 1 | Advanced |
| 265 | ro5185426.mp,kw. | 2 | Advanced |
| 266 | zelboraf*.mp,kw. | 42 | Advanced |
| 267 | 207smy3fqt.rn. | 1447 | Advanced |
| 268 | 918504-65-1.rn. | 0 | Advanced |
| 269 | or/75-268 | 177838 | Advanced |
| 270 | 74 and 269 | 7293 | Advanced |
| 271 | exp animals/ not (exp animals/ and exp humans/) | 4884720 | Advanced |
| 272 | 270 not 271 | 6964 | Advanced |
| 273 | limit 272 to yr="2010 -Current" | 6338 | Advanced |
